# Predicting suicide attempts in people with autism and co-morbid mood or psychotic disorders

**DOI:** 10.64898/2026.09.25.26364048

**Authors:** Monika Baker, Wai-Yin Lam, Mihai Virtosu, Nina de Lacy

**Affiliations:** Department of Psychiatry, University of Utah; Data Science Services, University of Utah; Huntsman Mental Health Institute

**Keywords:** Suicide, Attempted, Predictive Learning Models, Mood Disorders, Psychotic Disorders, Autism Spectrum Disorder

## Abstract

Individuals with autism spectrum disorder (ASD) and co-occurring mood or psychotic disorders face substantially elevated suicide-attempt risk, but prior research is largely limited to population-level incidence and cross-sectional risk-factor studies, leaving unclear whether individual-level, prospective prediction is possible in this population. We conducted a retrospective prediction model study using Epic Cosmos electronic health record (EHR) data from 322,879 individuals with ASD and co-occurring mood or psychotic disorders to predict suicide attempts at the next clinical encounter using only information available at or before the preceding encounter. Five machine-learning algorithms were compared, with performance evaluated across discrimination, calibration, and clinical utility. All algorithms achieved strong discrimination using the full predictor set (AUROC 0.87–0.91). A parsimonious feature set (22 predictors) matched or exceeded full-model performance, with extreme gradient boosting (XGBoost) achieving the highest discrimination and standardized net benefit post-ablation (AUROC = 0.90; AUPRC = 0.34), though net benefit was comparable for elastic net and logistic regression. A prior suicide attempt was the strongest individual predictor. Decision-curve analysis indicated that model-guided decisions provided greater net benefit than treating all or no individuals across a broad range of clinically plausible thresholds. These findings demonstrate that near-term suicide-attempt risk can be predicted with high decision utility in this high-risk population using data already captured in routine care, suggesting a feasible basis for future EHR-based clinical decision-support tools.

**Lay Summary:** People with autism spectrum disorder who also have a co-occurring mood disorder (such as depression or bipolar disorder) or a psychotic disorder face a higher risk of suicide attempts, but clinicians have had few tools to identify which patients are at greatest risk before an attempt occurs. Using medical records from over 320,000 people, we built a predictive model that estimates future risk of suicide attempt, using information already documented in routine care. The model performed strongly and points toward a practical way to help clinicians identify at-risk patients and intervene earlier.

## 1 Introduction

Individuals with autism spectrum disorder (ASD) face substantially elevated suicide-attempt risk (Richa et al., 2014; Segers & Rawana, 2014; Zahid & Upthegrove, 2017), yet clinicians currently have few tools to identify which individuals are at greatest near-term risk before an attempt occurs. Population-based studies in Sweden, Denmark, and Canada have shown that individuals with ASD are at greater risk of suicide attempts than individuals without ASD (Hirvikoski et al., 2020; Kõlves et al., 2021; Singal et al., 2026). In a nationwide Danish cohort of more than 6.5 million individuals, the adjusted incidence rate of suicide attempts was 3.19 times higher among individuals with ASD than among those without, with elevated rates observed across every age group examined (Kõlves et al., 2021).

This risk is further concentrated among individuals with co-occurring psychiatric conditions, particularly depressive, mood, and psychotic disorders (Hand et al., 2020; Kim et al., 2024; Kõlves et al., 2021): in the Danish cohort above, suicide-attempt incidence was 979 per 100,000 person-years among individuals with both ASD and an affective disorder and 736 per 100,000 person-years among those with both ASD and a schizophrenia spectrum disorder, both well above the overall incidence of 267 per 100,000 person-years among individuals with ASD (Kõlves et al., 2021). Mood and psychotic disorders confer similarly elevated suicide-attempt risk outside ASD-specific populations (Aaltonen et al., 2020; Fuller-Thomson & Hollister, 2016; Lu et al., 2020; Nock et al., 2009, 2010; Pallaskorpi et al., 2017), underscoring that individuals with ASD and co-occurring mood or psychotic disorders represent a population at especially compounded suicide-attempt risk.

Whether this risk can be identified prospectively at the individual level, however, remains unclear. Studies have reported associations between suicide risk and co-occurring psychiatric conditions, cognitive and emotional factors, and social or environmental stressors (Brown et al., 2024). However, this literature often combines suicidal ideation, attempts, and mortality and most studies are cross-sectional, limiting our ability to make specific prospective predictions about attempts (Brown et al., 2024; Kim et al., 2024). Beyond depressive disorders, attempt-specific evidence for other psychiatric diagnoses remains especially limited (Kim et al., 2024). A national Medicare claims study associated younger age, depression, intellectual disability, and greater psychiatric healthcare utilization with attempt- or self-injury-coded encounters among adults with ASD, but analyzed outcomes cross-sectionally and could not distinguish suicidal from non-suicidal self-injury (Hand et al., 2020). A cross-sectional machine-learning study similarly identified emotional and behavioral symptoms, family history, bullying, and routine changes as correlates of suicide-attempt status in ASD but classified existing status rather than predicting future attempts (Gharaibeh et al., 2026).

Studies in broader healthcare populations have shown that longitudinal electronic healthcare record (EHR) data can be used to predict future suicide attempts (Barak-Corren et al., 2017; Simon et al., 2018). Such predictions could help clinicians and health systems direct proactive follow-up and suicide-prevention resources toward individuals at greatest near-term risk (Barak-Corren et al., 2017; Hsin et al., 2026). However, prior work has not established whether longitudinal EHR data can prospectively identify individuals with ASD — particularly those with co-occurring mood or psychotic disorders — at greatest near-term risk, leaving clinicians without a decision-support tool to guide proactive follow-up and prevention.

Epic Cosmos aggregates longitudinal EHR data from hundreds of participating healthcare organizations and includes a broad range of routinely collected clinical variables (Tarabichi et al., 2021). Using this resource, the present study analyzed data from 322,879 individuals with ASD and co-occurring mood or psychotic disorders to predict future suicide attempt within 30 days, using only precedent information. Five machine-learning algorithms were compared, with potential clinical utility evaluated using decision-curve analysis and performance examined at thresholds reflecting different clinical capacities. Explainable models were constructed to identify the clinical features that contributed most strongly to model predictions. Together, this design moves beyond population-level associations and cross-sectional classification to evaluate near-term, individual-level prediction of suicide attempts among individuals with ASD and co-occurring mood or psychotic disorders.

## 2 Materials and Methods

### 2.1 Study Design and Data Source

This retrospective prediction model study drew on de-identified, national-scale EHR data housed in Epic Cosmos (Epic Systems Corporation, 2024), a data warehouse pooling records from over 2,223 hospitals and 50,900 clinics spanning more than 310 million unique patients across the United States. Because participating health systems contribute data that is aggregated and deidentified at the source, the resulting population reflects substantial geographic and demographic diversity across inpatient, outpatient, and emergency care settings. A unique patient identifier links each individual’s encounters across constituent health systems, consolidating what would otherwise be fragmented inpatient and outpatient charts into a single longitudinal record.

Before any site’s data enters Cosmos, local values are mapped to standardized vocabularies and transmitted securely; Epic then performs its own validation against source systems, correcting mapping discrepancies and evaluating completeness prior to approving the data for inclusion — a multi-stage curation process jointly overseen by Epic and each contributing health system. All analyses for this study were carried out entirely within the Cosmos Data Science Virtual Machine (DSVM), an environment that does not permit line-level data to leave its boundaries. The cohort was drawn from the period spanning January 1, 2016 through December 31, 2025.

### 2.2 Study cohort

Eligible individuals were required to have at least one clinical encounter between 2016 and 2025, a lifetime diagnosis of autism spectrum disorder (ASD), and a diagnosis of a mood disorder, psychotic disorder, or both. Diagnoses were identified from encounter-level International Classification of Diseases, Tenth Revision, Clinical Modification (ICD-10-CM) codes (National Center for Health Statistics, 2025). Mood and psychotic disorders were defined using the prespecified code set presented in Supplementary Table S1. ASD diagnoses were identified using the following ICD-10-CM codes: F84.0 (Autistic disorder), F84.5 (Asperger’s syndrome), F84.8 (Other pervasive developmental disorders), and F84.9 (Pervasive developmental disorder, unspecified). These eligibility criteria produced an initial cohort of 596,919 individuals before application of the encounter-level requirements described below.

### 2.3 Outcome Definition

The primary outcome was a recorded suicide attempt during the study period, identified using ICD-10-CM codes for intentional self-harm (Supplementary Table S2). This code set corresponds to the Centers for Disease Control and Prevention’s surveillance case definition for nonfatal suicide attempts (Hedegaard et al., 2018), the standard definition used in administrative-data suicide research. R45.88 (self-harm without suicidal intent), the ICD-10-CM code specifically designated to distinguish non-suicidal self-injury from suicidal self-harm, was not included in this code set. The outcome reported here therefore reflects recorded EHR-coded suicide attempt/self-harm events rather than confirmed suicide attempts verified by clinical or forensic review.

### 2.4 Temporally ordered point prediction

A temporally ordered prediction framework was used to prevent information recorded after the outcome from entering the predictor set, which could introduce temporal leakage and inflate apparent model performance (Yuan et al., 2021). One encounter per individual was designated as the index encounter and used to determine suicide-attempt status. The immediately preceding eligible encounter was designated as the feature encounter. Only information available at or before the feature encounter contributed to model predictors; no information from the index encounter was included among the features. Individuals were required to have at least two encounters: one feature encounter and one index encounter. The feature encounter was required to occur more than 0 and no more than 30 days before the index encounter, and age at the index encounter was restricted to 10–100 years. For each individual, the most recent available encounter was first considered as the index encounter and the preceding encounter as the feature encounter. If the encounter pair did not satisfy the eligibility requirements, the index encounter was moved to the next most recent encounter and eligibility was reassessed. This process continued until an eligible pair was identified or fewer than two encounters remained, in which case the individual was excluded. The target was coded as positive when a suicide attempt occurred at the resulting index encounter and negative otherwise. Application of these criteria to the initial cohort of 596,919 individuals yielded a final sample of 322,879 individuals eligible for point prediction.

### 2.5 Features

Candidate predictors comprised patient-level and encounter-level EHR variables. Patient-level variables represented characteristics that were fixed or changed slowly over time and were not tied to a specific encounter, including demographic and socioeconomic characteristics.

Encounter-level variables represented clinical characteristics documented at the feature encounter or during the period preceding it, including age, anthropometric measures, diagnoses, and healthcare utilization. For point prediction, the patient-level and feature-encounter variables were combined into a single fixed-length vector for each individual. A complete list of candidate variables and their definitions is provided in Supplementary Table S3.

### 2.6 Data cleaning and preprocessing

Variables categorizing the self-reported race of each individual and other nominal variables were one-hot encoded. Patient-level dates were represented as the number of days since birth, and encounter-level dates were represented as the number of days preceding the feature encounter. ICD-10-CM diagnosis variables were grouped into the 22 ICD-10-CM chapters and one-hot encoded. After filtering and encoding, 123 patient level-features and 91 encounter-level features remained for modeling. Individuals were randomly divided into training and out of sample held-out test sets in an 80:20 ratio, stratified by the outcome. All subsequent preprocessing was performed separately within the training and test sets. Continuous variables were clipped at the training-set mean plus or minus three standard deviations, ordinal variables were clipped to ranges specified in the data dictionary, and variables were scaled to the interval [0, 1] using min-max normalization. Negative values in derived date variables were replaced with missing values to reflect data quality issues.

Missingness for patient-level variables was assessed across individuals, whereas missingness for encounter-level variables was assessed across feature encounters. Cosmos distinguishes between some forms of missingness, such as refusal to answer. These values were represented using binary missingness indicators; the remaining missing values were treated as missing at random. Features with more than 35% missingness were excluded from subsequent analysis. The primary analysis used zero imputation accompanied by a binary indicator for each imputed feature. Simple rather than model-based imputation was used because this is the conventional way of imputing missing values in models intended for use as clinical decision support tools that require filling missing values at run time deployment. Because missingness indicators can retain predictive information contained in missing-data patterns (Sisk et al., 2023), a sensitivity analysis comparing zero, mean, and median imputation is presented in Supplementary Table S4.

### 2.7 Model development

Five point-prediction algorithms were evaluated: logistic regression (LR), elastic-net logistic regression (EN), linear support vector machine (SVM), extreme gradient boosting (XGBoost), and an artificial neural network (ANN). Models were implemented in Python. LR, EN, and SVM were developed using scikit-learn (Pedregosa et al., 2011); XGBoost used the XGBoost library (Chen & Guestrin, 2016); and the ANN was implemented in PyTorch (Paszke et al., 2019).

#### 2.7.1 Standard Learning

For all five algorithms, hyperparameters were selected for reporting via grid search under 5-fold stratified cross-validation on the training set, using AUPRC as the tuning criterion, given its suitability for imbalanced classification tasks (Saito & Rehmsmeier, 2015). The winning configuration for each algorithm was refit on an 80%/20% stratified split of the training data, with the 80% subset used for final model fitting and the 20% held out for post-hoc probability calibration (Platt scaling, isotonic regression, and temperature scaling).

#### 2.7.2 Cost-sensitive learning

As a sensitivity analysis, each of the five algorithms was refit under cost-sensitive learning (CSL), which assigns asymmetric weights to false positives and false negatives during training rather than treating both error types as equally costly, shifting the model’s decision boundary toward a pre-specified operating region (Elkan, 2001). CSL models followed the same grid-search and cross-validation procedure as the standard-learning models described above, with the false-positive-averse cost multiplier added to each algorithm’s hyperparameter grid and tuned jointly with its other hyperparameters, using recall at the 99th percentile of specificity as the tuning criterion to enforce the pre-specified low-false-positive operating region (Wynants et al., 2019). CSL models are reported in Supplementary Table S6.

### 2.8 Model evaluation

Model performance was evaluated using complementary measures of discrimination, classification performance, calibration, and clinical utility, consistent with the STRATOS framework and PROBAST+AI guidance (Van Calster et al., 2025; Moons et al., 2025). Discrimination was quantified using the area under the receiver operating characteristic curve (AUROC) and AUPRC. Threshold-dependent measures included positive predictive value (PPV), sensitivity, specificity, and Youden’s index, calculated as sensitivity plus specificity minus 1 and balanced accuracy, calculated as the mean of sensitivity and specificity. Performance was evaluated at the default probability threshold of 0.50 and at several prespecified operating points. Thresholds were also set at the 90th, 95th, and 99th percentiles of predicted risk among test-set individuals without a suicide attempt, fixing specificity at each of these levels. Separately, individuals were ranked by predicted risk and the highest-risk 1%, 2%, and 5% were classified as positive. Because these top-risk thresholds are defined by rank rather than a probability cutoff, they flagged the same number of individuals across all models.

Calibration was evaluated with four measures: the calibration intercept and slope, expected calibration error (ECE), and Brier score. Intercept and slope came from regressing the observed outcome on the logit of predicted risk, where 0 and 1 mark perfect calibration, respectively. For ECE, predicted probabilities were split into deciles, and we took the mean absolute difference between predicted and observed risk across those bins. Brier score is the mean squared difference between predicted probabilities and observed outcomes.

Decision-curve analysis was used to evaluate standardized net benefit across threshold probabilities (Vickers & Elkin, 2006). Net benefit was calculated as (TP/N) - (FP/N) x [p_t_/(1 - p_t_)], where TP and FP denote true- and false-positive classifications, N is the test-set sample size, and p_t_ is the threshold probability. Standardized net benefit was obtained by dividing net benefit by the observed outcome prevalence. A threshold probability of approximately 9% represented a 10:1 relative weighting of a missed attempt compared with an unnecessary clinical assessment. Results were also presented across a range of threshold probabilities rather than interpreted at a single threshold.

### 2.9 Feature ablation

Feature ablation was performed to construct the performance of more parsimonious models, since these offer speed performances at run time. Feature importance was defined as the absolute fitted coefficient for LR, EN, and SVM, and via SHAP (SHapley Additive exPlanations) for the tree-based and neural network models. SHAP is a game-theoretic framework that attributes each feature a contribution to a given prediction based on its Shapley value — its average marginal effect on the model’s output for a given participant across all possible orderings in which features could be added (Lundberg & Lee, 2017). SHAP values were computed per participant and per feature: for XGBoost using the XGBoost library’s native SHAP implementation, and for the ANN via GradientSHAP, a gradient-based SHAP approximation, using Captum (Kokhlikyan et al., 2020). Global feature importance was then defined, for both algorithms, as the mean absolute SHAP value averaged across participants.

We then used a principled method, adapted from the feature ablation approach implemented in RiskPath (de Lacy et al., 2024), to determine the number and identity of features in parsimonious models where adding additional features offered diminishing returns. Predictors were ranked in decreasing order of importance, and models were refitted using progressively larger subsets of the highest-ranked predictors. The retained feature set was defined using a knee point in the importance-rank curve, identified as the point of maximum perpendicular distance from the chord connecting the curve’s endpoints, based on the method implemented in the *kneefinder* package (Lavorini, 2021). SHAP beeswarm plots were used to display the direction and magnitude of feature contributions for models with SHAP-based importance estimates.

### 2.10 Ethics

This study was determined to not meet the definitions of Human Subjects Research by the University of Utah Institutional Review Board (IRB_00202177) and was therefore exempt from informed consent requirements.

### 2.11 Data Availability Statement

The data used in this study are derived from the Epic Cosmos data warehouse and are not publicly available due to data privacy and security restrictions inherent to the Cosmos Data Science Virtual Machine (DSVM) environment. Researchers interested in accessing Epic Cosmos data should contact Epic Systems or their participating institution. The analytic code used in this study is available from the de Lacy lab’s GitHub (https://github.com/delacylab/Cosmos_Modeling).

## 3 Results

### 3.1 Sample Characteristics

The final study cohort comprised 322,879 individuals meeting study inclusion criteria, of whom 317,112 met criteria for a mood disorder, 84,364 for a psychotic disorder, and 78,597 for both conditions simultaneously (Table 1). The cohort was majority male, and the proportion of male individuals was modestly higher among those meeting psychosis criteria than in the overall sample. Race and ethnicity were predominantly White and non-Hispanic, respectively, though both fields — along with gender identity — showed appreciable rates of missing or unrecorded data (Table 1). Of 322,879 individuals, 3,143 (0.97%) had a recorded suicide attempt at the index encounter.

**Table 1.** Demographic Characteristics of Study Participants. Mood and Psychosis reflect patients meeting each criterion independently (not mutually exclusive); “Both” is the subset meeting both simultaneously.

| **Characteristic** | **All**<br>**(N=322,879)** | **Mood**<br>**(N=317,112)** | **Psychosis**<br>**(N=84,364)** | **Both**<br>**(N=78,597)** |
| --- | --- | --- | --- | --- |
| **Sex** |  |  |  |  |
| Female | 135,548 (42.0%) | 134,239 (42.3%) | 31,409 (37.2%) | 30,100 (38.3%) |
| Male | 186,548 (57.8%) | 182,092 (57.4%) | 52,791 (62.6%) | 48,335 (61.5%) |
| Other | 164 (0.1%) | 163 (0.1%) | 38 (0.0%) | 37 (0.0%) |
| Not recorded/missing | 619 (0.2%) | 618 (0.2%) | 126 (0.1%) | 125 (0.2%) |
| **Gender Identity** |  |  |  |  |
| Female | 55,588 (17.2%) | 55,081 (17.4%) | 13,501 (16.0%) | 12,994 (16.5%) |
| Male | 70,637 (21.9%) | 69,019 (21.8%) | 21,885 (25.9%) | 20,267 (25.8%) |
| Transgender male | 4,561 (1.4%) | 4,554 (1.4%) | 1,065 (1.3%) | 1,058 (1.3%) |
| Transgender female | 3,754 (1.2%) | 3,748 (1.2%) | 807 (1.0%) | 801 (1.0%) |
| Choose not to disclose | 306 (0.1%) | 302 (0.1%) | 73 (0.1%) | 69 (0.1%) |
| Not recorded/missing | 188,033 (58.2%) | 184,408 (58.2%) | 47,033 (55.8%) | 43,408 (55.2%) |
| **Race** |  |  |  |  |
| American Indian/Alaska Native | 1,100 (0.3%) | 1,075 (0.3%) | 312 (0.4%) | 287 (0.4%) |
| Asian | 3,586 (1.1%) | 3,440 (1.1%) | 1,093 (1.3%) | 947 (1.2%) |
| Black or African American | 23,037 (7.1%) | 21,933 (6.9%) | 9,974 (11.8%) | 8,870 (11.3%) |
| Native Hawaiian/Pacific Islander | 246 (0.1%) | 239 (0.1%) | 76 (0.1%) | 69 (0.1%) |
| Other Race | 7,505 (2.3%) | 7,285 (2.3%) | 2,215 (2.6%) | 1,995 (2.5%) |

| Characteristic | All<br>(N=322,879) | Mood<br>(N=317,112) | Psychosis<br>(N=84,364) | Both<br>(N=78,597) |
| --- | --- | --- | --- | --- |
| White | 205,349 (63.6%) | 202,671 (63.9%) | 48,624 (57.6%) | 45,946 (58.5%) |
| Multiple Races | 47,729 (14.8%) | 46,878 (14.8%) | 14,864 (17.6%) | 14,013 (17.8%) |
| Not recorded/missing | 34,327 (10.6%) | 33,591 (10.6%) | 7,206 (8.5%) | 6,470 (8.2%) |
| **Ethnicity** |  |  |  |  |
| Hispanic or Latino | 22,223 (6.9%) | 21,642 (6.8%) | 6,317 (7.5%) | 5,736 (7.3%) |
| Non-Hispanic | 252,051 (78.1%) | 247,810 (78.1%) | 66,303 (78.6%) | 62,062 (79.0%) |
| Missing | 48,605 (15.1%) | 47,660 (15.0%) | 11,744 (13.9%) | 10,799 (12.7%) |

### 3.2 Ensemble-based learning showed the strongest overall discrimination performance in predicting 30-day suicide attempts in people with autism

Across the five algorithms evaluated using this configuration and the full (pre-ablation) feature set, discrimination was consistently very strong, with AUROC ranging from 0.87 (ANN) to 0.91 (XGBoost) (Table 2a). Given the low outcome prevalence, AUPRC — a more informative measure of discrimination under class imbalance (Saito & Rehmsmeier, 2015) — showed greater separation between algorithms, with XGBoost achieving the highest value (0.25) and ANN the lowest (0.12). Calibration was generally strong across algorithms following isotonic calibration. Calibration slopes were nearest to the ideal value of 1 for the support vector machine (1.002) and XGBoost (0.999), while ANN, elastic net, and logistic regression showed comparable, modest attenuation (slopes ranging from 0.963 to 0.968). Following feature ablation, performance was similar to or better than the full-feature models despite substantial reductions in feature count (Table 2b). XGBoost showed the largest improvement, with AUPRC increasing from 0.252 to 0.336 using only 22 of the original features, alongside a marginal decrease in AUROC. Elastic net and the support vector machine showed comparable feature reductions (18 and 42 features, respectively) without meaningful loss of discrimination. Among the imputation and calibration strategies evaluated, zero imputation and isotonic calibration yielded the strongest performance and were adopted for the primary analysis.

**Table 2.** Model Performance Metrics (Zero Imputation, Isotonic Calibration) **a) Pre-ablation** All models use zero imputation, isotonic calibration, pre-ablation feature set, and standard learning. Recall@99Spec = recall at the threshold corresponding to 99th-percentile specificity. Precision@top1% = precision among the top 1% highest-risk patients. Balanced accuracy and standardized net benefit are evaluated at the policy decision threshold (p_t = 1/11), consistent with the net-benefit weighting used throughout. The best performance for each metric is highlighted in blue. **b) Post-ablation** All models use zero imputation, isotonic calibration, pre-ablation feature set, and standard learning. Recall@99Spec = recall at the threshold corresponding to 99th-percentile specificity. Precision@top1% = precision among the top 1% highest-risk patients. Balanced accuracy and standardized net benefit are evaluated at the policy decision threshold (p_t = 1/11), consistent with the net-benefit weighting used throughout. Features = number of retained predictors after ablation (varies by algorithm, selected using ablation method).

| Algorithm | AUROC | AUPRC | Recall@99Spec | Precision@top1% | Calibration Slope | Calibration Intercept | Balanced Accuracy | Std. Net Benefit |
| --- | --- | --- | --- | --- | --- | --- | --- | --- |
| ANN | 0.8707 | 0.1221 | 0.2513 | 0.2157 | 0.9675 | -0.0415 | 0.6832 | 0.1760 |
| EN | 0.9013 | 0.2073 | 0.4920 | 0.3478 | 0.9626 | -0.0210 | 0.7411 | 0.3928 |
| LogReg | 0.9051 | 0.2291 | 0.4592 | 0.3602 | 0.9674 | -0.0392 | 0.7374 | 0.3759 |
| SVM | 0.9008 | 0.2249 | 0.4857 | 0.3540 | 1.0022 | -0.0301 | 0.7465 | 0.3859 |
| XGB | 0.9116 | 0.2521 | 0.4825 | 0.3715 | 0.9999 | -0.0273 | 0.7429 | 0.3897 |

| Algorithm | Features | AUROC | AUPRC | Recall@99Spec | Precision@top1% | Calibration Slope | Calibration Intercept | Balanced Accuracy | Std. Net Benefit |
| --- | --- | --- | --- | --- | --- | --- | --- | --- | --- |
| ANN | 61 | 0.8605 | 0.1277 | 0.2078 | 0.2188 | 1.0061 | -0.0252 | 0.6410 | 0.1638 |
| EN | 18 | 0.8988 | 0.1976 | 0.4899 | 0.3488 | 0.9887 | -0.0174 | 0.7403 | 0.3942 |
| LogReg | 37 | 0.8663 | 0.2121 | 0.4867 | 0.3529 | 0.9565 | -0.0204 | 0.7382 | 0.3915 |
| SVM | 42 | 0.8464 | 0.2376 | 0.4889 | 0.3498 | 0.9842 | -0.0090 | 0.7320 | 0.3823 |
| XGB | 22 | 0.9027 | 0.3360 | 0.4920 | 0.3664 | 0.9975 | -0.0208 | 0.7412 | 0.3943 |

The full set of performance metrics, including additional threshold-dependent and calibration measures beyond those summarized here, is reported in Supplementary Table S5. Sensitivity analyses examining alternative imputation strategies (Supplementary Table S4), cost-sensitive learning (Supplementary Table S6), and alternative calibration methods (Supplementary Table S8) are presented for comparison. Under cost-sensitive learning, EN, logistic regression, and SVM showed performance nearly identical to their standard learning counterparts, while the ANN and XGBoost models showed substantial degradation in discrimination (AUROC 0.87→0.70 and 0.91→0.74, respectively), suggesting these two algorithms were comparatively unstable under the asymmetric class reweighting evaluated here.

### 3.3 Clinical predictors were the most important contributors to performance

Figure 1 presents SHAP values for the ten predictors contributing most strongly, on average, to individual risk predictions in the post-ablation XGBoost model. XGBoost was selected as the representative model on the basis of its discrimination, calibration, and standardized net benefit, each among the highest observed across the five algorithms (Table 2b); standardized net benefit, in particular, was nearly identical for elastic net and logistic regression, indicating comparable clinical utility among these top-performing algorithms. Corresponding beeswarm plots for the pre-ablation XGBoost model and for the ANN model (pre- and post-ablation) are provided for comparison in Supplementary Figure S1a–c. For most binary predictors, presence of the feature was associated with higher predicted risk: a recorded suicide attempt at the feature encounter, prior suicide attempts, substance use disorder diagnoses, personality disorder diagnoses, and missingness of primary infections diagnosis data each showed higher SHAP values when present and lower SHAP values when absent. Two diagnostic-category variables showed the inverse pattern, with feature presence associated with lower predicted risk: a non-primary psychiatric diagnosis and a primary psychiatric diagnosis in the three months preceding the feature encounter. Number of ED visits in the preceding three months showed a distinct, threshold-like pattern: low utilization was associated with a wide range of SHAP values, while the highest predicted-risk contributions were concentrated among patients with the greatest number of ED visits. Age at the feature encounter was inversely associated with risk, with younger age corresponding to higher SHAP values and older age corresponding to lower, protective SHAP values across a broad range; the age at which alcohol use history was recorded showed a similar protective pattern.

**Figure 1.**
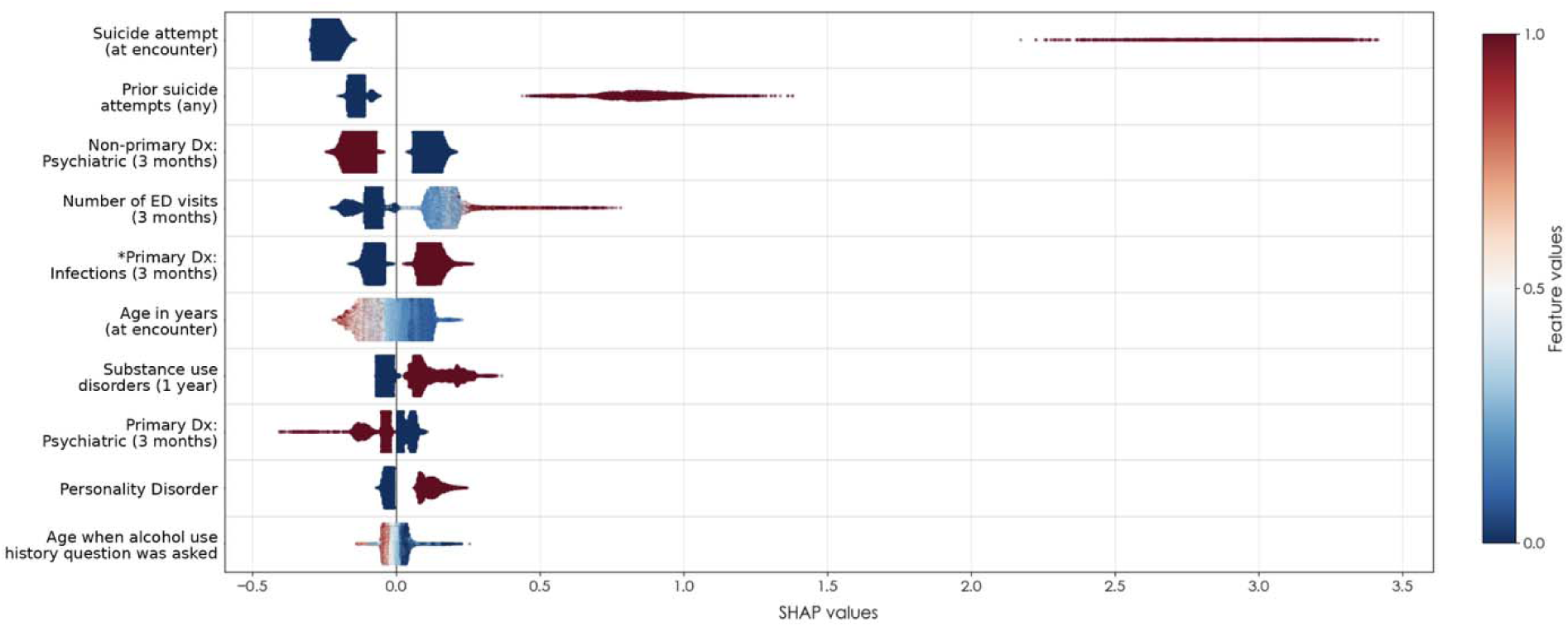
Beeswarm plot of top features for XGBoost model, post-ablation. Each point represents an individual in the held-out test set; color reflects the min-max normalized feature value (blue = lower, red = higher), and horizontal position reflects each feature’s contribution to that individual’s predicted risk. Features are ordered by mean absolute SHAP value. Asterisked features are binary missingness indicators. Dx = diagnosis; ED = emergency department. See Supplementary Table 3 for full feature definitions.

### 3.4 Strong decision utility in predicting suicide was obtained across models

Decision-curve analysis showed that all five algorithms provided greater standardized net benefit than strategies of treating all or no individuals across most of the threshold range examined, with net benefit converging toward that of treating no individuals near a threshold probability of 35–39% (Figure 2). Elastic net, logistic regression, support vector machine, and XGBoost performed comparably across nearly the entire threshold range, with largely overlapping curves despite the differences in discrimination reported above. ANN provided the lowest net benefit of the five algorithms at every threshold examined, consistent with its comparatively lower discrimination (Table 2a). At a threshold probability of approximately 9%, corresponding to weighting a missed suicide attempt ten times more heavily than an unnecessary clinical assessment, all algorithms except ANN offered substantially greater net benefit than either treat-all or treat-none strategies.

**Figure 2.**
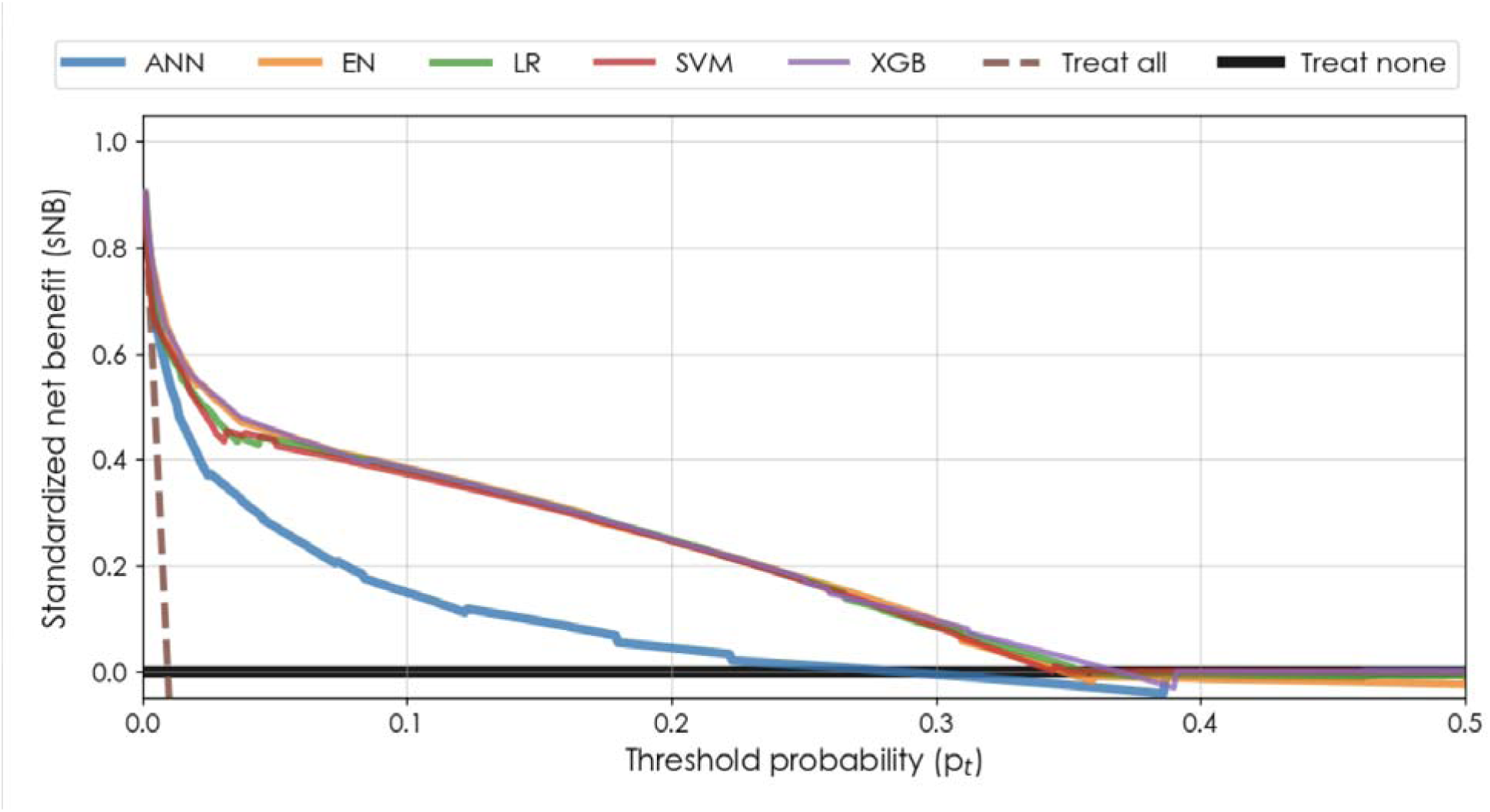
Decision Curve Analysis for point prediction models. Standardized net benefit (sNB) is shown as a function of the decision threshold probabilities (p_t_) for five point prediction models (post-ablation) evaluated on the same held-out test set. Features were zero-imputed with additional binary indication for data missingness. The best performance achieved from standard learning is reported. The x-axis is limited to p_t_ ≤ 0.5 to preserve legibility at higher threshold values. The treat-all strategy (brown dashed line) represents the sNB obtained by assigning the intervention to all individuals regardless of predicted risk, while the treat-none strategy (solid black horizontal line) corresponds to withholding intervention from all individuals. ANN = Artificial Neural Network; EN = Elastic Net; LG = Logistic Regression; SVM = Support Vector Machine; XGB = Extreme Gradient Boosting.

## Discussion

### 4.1 Principal Findings

In this large, retrospective prediction model study of 322,879 individuals with autism spectrum disorder and co-occurring mood or psychotic disorders, five machine-learning algorithms demonstrated strong and broadly comparable discrimination for predicting a near-term suicide attempt using routinely collected EHR data. Using a substantially reduced, SHAP-selected feature set (22 of the original predictors), XGBoost achieved the highest discrimination and standardized net benefit (AUROC = 0.90; AUPRC = 0.34), though elastic net and logistic regression achieved nearly identical standardized net benefit, indicating comparable clinical utility across these top-performing algorithms. This reduced feature set performed as well as or better than the full predictor set for XGBoost, indicating that near-term risk can be captured with a comparatively parsimonious model. A recorded suicide attempt at the preceding encounter was the strongest individual predictor, with several markers of psychiatric acuity and healthcare utilization contributing additional, smaller effects. Decision-curve analysis indicated that model-guided decisions offered greater net benefit than treating all or no individuals across threshold probabilities up to approximately 35–39%, beyond which net benefit converged toward that of treating no individuals.

### 4.2 Comparison with Prior Literature

The strong discrimination observed across all five algorithms, and the clinical utility demonstrated through decision-curve analysis, indicate that near-term suicide-attempt risk can be predicted with meaningful accuracy in this population using routinely collected EHR data. This extends a body of work showing that EHR-based prediction is feasible in general clinical populations (Barak-Corren et al., 2017; Simon et al., 2018) to a population whose elevated risk has previously been characterized only through population-level incidence and cross-sectional association (Hand et al., 2020; Kõlves et al., 2021). Unlike the only prior machine-learning application specific to suicide attempts in ASD, which classified existing attempt status rather than forecasting subsequent risk (Gharaibeh et al., 2026), the temporally ordered design used here supports genuine prospective prediction.

### 4.3 Feature Importance

The two strongest predictors identified — a recorded suicide attempt at the feature encounter and a history of prior suicide attempts — align closely with an extensive general-population literature identifying prior suicidal behavior as among the most robust known predictors of future attempts (De la Torre-Luque et al., 2023). This pattern has also been specifically documented among individuals with ASD, where suicide-attempt risk is markedly elevated relative to the general population (Kõlves et al., 2021), and among individuals with mood and psychotic disorders, where attempts have been shown to cluster during periods of acute symptom exacerbation (Aaltonen et al., 2020; Pallaskorpi et al., 2017). The convergence of this predictor’s importance across general, ASD-specific, and mood/psychosis-comorbid populations suggests that recent suicidal behavior functions as a consistent, transdiagnostic risk signal, one that retains strong predictive value even within a population already selected for elevated baseline risk.

Substance use disorder and personality disorder diagnoses were also associated with higher predicted risk, consistent with well-established general-population evidence linking both conditions to elevated suicide-attempt risk (McClelland et al., 2023; Wilcox et al., 2004). Within ASD specifically, substance use disorders have similarly been shown to confer substantially elevated suicide-attempt risk, with certain subtypes conferring particularly large relative increases (Baker et al., 2026). Notably, a variable specific to borderline personality disorder was not among the ten most influential predictors, despite borderline personality disorder carrying the strongest documented suicide-risk association among personality disorders in the general population; this dissociation may indicate that the broader personality disorder category captured signal not specific to borderline disorder pathology.

Number of emergency department (ED) visits showed a distinct, non-monotonic relationship with predicted risk: at low utilization, SHAP values spanned a wide range, whereas the highest predicted-risk contributions were concentrated among individuals with the greatest number of ED visits. This pattern is consistent with prior evidence that ED visit frequency is independently associated with suicide risk in a dose-dependent manner, with risk rising sharply among individuals with four or more visits (Kvaran et al., 2015).

Age at the feature encounter was inversely associated with predicted risk, with younger individuals showing higher SHAP values across a broad range. This pattern is consistent with cross-national epidemiological evidence identifying younger age as one of the most consistent risk factors specifically for suicide attempts, alongside female sex, lower educational attainment, and co-occurring psychiatric illness (Nock et al., 2008). A related but distinct pattern was observed for the age at which alcohol use history was documented, which showed a similar inverse gradient over a narrower range. Because detection of alcohol-related concerns in clinical practice often relies on clinician suspicion rather than routine, standardized screening (Vinson et al., 2013), this variable may function less as a measure of alcohol use itself and more as a marker of the age at which clinical concern prompted inquiry — though this interpretation remains speculative given the absence of direct evidence for this specific variable.

Two diagnostic-category variables showed an inverse relationship with predicted risk: presence of a non-primary or primary psychiatric diagnosis in the preceding months was associated with lower, rather than higher, predicted risk. This pattern may reflect the role of treatment engagement and continuity of psychiatric care, which has been independently associated with reduced suicide risk following psychiatric contact (Choi et al., 2020), including specifically among patients with mood disorders, for whom greater continuity of care has been associated with lower suicide mortality (Kim et al., 2018). Under this interpretation, a documented psychiatric visit may function as a marker of active clinical contact and ongoing care rather than acute psychiatric crisis, though this study cannot distinguish between these explanations directly.

### 4.4 Clinical Implications

The decision-curve and feature-ablation results together suggest concrete ways this model could inform near-term clinical decision-making in this population. Decision-curve analysis indicated that model-guided decisions strong potential clinical utility. This range spans clinically relevant tradeoffs between the relative costs of false positives and false negatives, meaning the model retained net benefit whether a given clinical setting prioritized minimizing missed attempts, accepting more unnecessary assessments to do so, or prioritized limiting those assessments and acting only on higher-confidence predictions. In practice, this allows the operating threshold to be selected according to locally available resources for follow-up and outreach, rather than requiring a single universal cutoff, while still outperforming a strategy of treating all or no individuals at any threshold within this range. This mirrors the general approach used by the Veterans Health Administration’s REACH VET program, which flags the highest-risk patients each month for proactive outreach, safety planning, and care re-engagement; implementation of that program has been associated with increased outpatient follow-up and reduced emergency department visits and suicide attempts among flagged patients (McCarthy et al., 2015, 2021). A comparable workflow could be implemented for individuals with ASD and co-occurring mood or psychotic disorders: following a qualifying outpatient encounter, patients in the highest-risk percentile could be automatically flagged for care coordinator outreach, reassessment of safety planning using an established protocol (Stanley & Brown, 2012), or brief follow-up contact, an intervention independently associated with reduced suicide risk following identification of elevated risk (Motto & Bostrom, 2001).

The feature ablation results further suggest that such a tool would not necessarily require extensive data collection to be useful. A 22-feature model achieved discrimination comparable to or exceeding the full model, drawing on variables already routinely documented in clinical care, including recent suicide-related history, healthcare utilization, and common psychiatric and substance use diagnoses. This reduces the practical burden of implementing risk stratification within existing EHR infrastructure, in contrast to approaches that depend on specialized instruments or active patient self-report not routinely captured in usual care (Bentley et al., 2022).

### 4.5 Strengths and Limitations

This study has several notable strengths. First, the analytic sample was large and drawn from a national EHR data warehouse spanning hundreds of participating health systems, supporting a level of demographic and geographic diversity not typically available in single-site studies. Second, the temporally ordered prediction framework restricted predictors to information available before the index encounter, addressing a limitation common to much of the existing ASD-specific suicide-risk literature, which has relied primarily on cross-sectional or concurrent classification approaches. Third, model development drew on a broad candidate feature set spanning 214 patient- and encounter-level variables, and incorporated a systematic comparison across five algorithms and multiple imputation strategies rather than a single default pipeline, strengthening confidence that the reported performance reflects genuine predictive signal rather than an artifact of a particular modeling choice. Fourth, feature ablation demonstrated that a substantially reduced predictor set could match or exceed full-model performance, supporting the feasibility of a more parsimonious, clinically translatable model. Finally, model evaluation extended beyond discrimination to include calibration and clinical utility via decision-curve analysis, providing a more complete picture of model performance than discrimination alone would offer.

This study also has several limitations. The outcome was ascertained using ICD-10-CM diagnosis codes for intentional self-harm rather than through direct clinical confirmation of each suicide attempt, and although this code set corresponds to the CDC’s surveillance case definition for nonfatal suicide attempts, administrative coding for suicide attempts has been shown to have limited sensitivity and variable positive predictive value across health systems, meaning both missed and misclassified events are possible (Hensley et al., 2026). A related limitation concerns the low outcome prevalence: even a well-discriminating model can yield a modest positive predictive value at low base rates, meaning that a substantial proportion of flagged individuals at any given threshold will not go on to have a suicide attempt, a constraint inherent to suicide-attempt prediction broadly rather than specific to this model (Belsher et al., 2019). Application of the study’s encounter-pairing and timing criteria reduced the initial cohort of 596,919 individuals to a final sample of 322,879; because inclusion required a qualifying pair of encounters, individuals with less frequent healthcare contact were systematically less likely to be captured, a form of bias well documented in EHR-derived research (Goldstein et al., 2016). Several demographic variables, including race, ethnicity, and gender identity, had substantial missingness (Table 1); missingness in race and ethnicity fields in EHR data has been shown to be frequently nonrandom rather than missing at random, which may limit the interpretability of demographic patterns reported here (Grundmeier et al., 2015). The point-prediction framework evaluated a single feature-index encounter pair per individual rather than modeling risk continuously over time, an approach that has been argued to be inherently limited relative to dynamic, continuously updated risk prediction (Nguyen et al., 2025). Finally, although Cosmos aggregates data from a large, diverse set of participating health systems, model performance was evaluated on a held-out test set drawn from the same data source and time period as model development; external validation in independent cohorts and healthcare contexts is necessary before generalizability can be assumed (Steyerberg & Harrell, 2016).

### 4.6 Conclusion

In a large, national sample of individuals with autism spectrum disorder and co-occurring mood or psychotic disorders, machine-learning models trained on routinely collected electronic health record data achieved strong discrimination and clinical utility for predicting near-term suicide attempts, with a substantially reduced feature set matching or exceeding full-model performance. These findings extend existing population-level and cross-sectional evidence of elevated suicide-attempt risk in this population toward individual-level, prospective risk stratification, and suggest that this model could support clinical decision-making, such as flagging patients for proactive outreach and safety planning, using data already captured in usual care rather than specialized screening instruments. Future work will evaluate whether modeling longitudinal patterns across varying numbers of encounters and lookback windows, rather than a single feature-index encounter pair, further improves prediction in this population (de Lacy et al., 2025).

## Supporting information

Data Dictionary

## Data Availability

The data used in this study are derived from the Epic Cosmos data warehouse and are not publicly available due to data privacy and security restrictions inherent to the Cosmos Data Science Virtual Machine (DSVM) environment. Researchers interested in accessing Epic Cosmos data should contact Epic Systems or their participating institution. The analytic code used in this study is available from the de Lacy labs GitHub (https://github.com/delacylab/Cosmos_Modeling).

https://cosmos.epic.com

## Acknowledgments

The authors would like to thank David Danks for his contributions to the content of the methodology and the manuscript. The authors acknowledge the use of Claude (Anthropic, San Franciso, CA) for assistance with language editing and manuscript revision. All content was reviewed, verified, and approved by the authors, who take responsibility for the accuracy of the published work. The senior author received financial support from the Huntsman Mental Health Foundation.

## Notes

### Competing Interest Statement

The authors have declared no competing interest.

