## Supplemental Tables for "Predicting suicide attempts in people with autism and co-morbid mood or psychotic disorders"

SUPPLEMENTARY TABLE S1 | ICD-10-CM diagnosis codes used to define mood and psychotic disorders.

Codes were used to identify individuals with a qualifying mood disorder, psychotic disorder, or both for inclusion in the study cohort.

| **Diagnostic category** | **ICD-10-CM code** | **Diagnosis** |
| --- | --- | --- |
| Psychotic disorder | F20.0 | Paranoid schizophrenia |
| Psychotic disorder | F20.1 | Disorganized schizophrenia |
| Psychotic disorder | F20.2 | Catatonic schizophrenia |
| Psychotic disorder | F20.3 | Undifferentiated schizophrenia |
| Psychotic disorder | F20.5 | Residual schizophrenia |
| Psychotic disorder | F20.81 | Schizophreniform disorder |
| Psychotic disorder | F20.89 | Other schizophrenia |
| Psychotic disorder | F20.9 | Schizophrenia, unspecified |
| Psychotic disorder | F21 | Schizotypal disorder |
| Psychotic disorder | F22 | Delusional disorders |
| Psychotic disorder | F23 | Brief psychotic disorder |
| Psychotic disorder | F24 | Shared psychotic disorder |
| Mood and psychotic disorders | F25.0 | Schizoaffective disorder, bipolar type |
| Mood and psychotic disorders | F25.1 | Schizoaffective disorder, depressive type |
| Mood and psychotic disorders | F25.8 | Other schizoaffective disorders |
| Mood and psychotic disorders | F25.9 | Schizoaffective disorder, unspecified |
| Psychotic disorder | F28 | Other psychotic disorder not due to a substance or known physiological condition |
| Psychotic disorder | F29 | Unspecified psychosis not due to a substance or known physiological condition |
| Mood disorder | F30.10 | Manic episode without psychotic symptoms, unspecified |
| Mood disorder | F30.11 | Manic episode without psychotic symptoms, mild |
| Mood disorder | F30.12 | Manic episode without psychotic symptoms, moderate |
| Mood disorder | F30.13 | Manic episode, severe, without psychotic symptoms |
| Mood and psychotic disorders | F30.2 | Manic episode, severe with psychotic symptoms |
| Mood disorder | F30.3 | Manic episode in partial remission |
| Mood disorder | F30.4 | Manic episode in full remission |
| Mood disorder | F30.8 | Other manic episodes |
| Mood disorder | F30.9 | Manic episode, unspecified |
| Mood disorder | F31.0 | Bipolar disorder, current episode hypomanic |
| Mood disorder | F31.10 | Bipolar disorder, current episode manic without psychotic features, unspecified |
| Mood disorder | F31.11 | Bipolar disorder, current episode manic without psychotic features, mild |
| Mood disorder | F31.12 | Bipolar disorder, current episode manic without psychotic features, moderate |
| Mood disorder | F31.13 | Bipolar disorder, current episode manic without psychotic features, severe |
| Mood and psychotic disorders | F31.2 | Bipolar disorder, current episode manic, severe with psychotic features |
| Mood disorder | F31.30 | Bipolar disorder, current episode depressed, mild or moderate severity, unspecified |
| Mood disorder | F31.31 | Bipolar disorder, current episode depressed, mild |
| Mood disorder | F31.32 | Bipolar disorder, current episode depressed, moderate |
| Mood disorder | F31.4 | Bipolar disorder, current episode depressed, severe, without psychotic features |
| Mood and psychotic disorders | F31.5 | Bipolar disorder, current episode depressed, severe with psychotic features |
| Mood disorder | F31.60 | Bipolar disorder, current episode mixed, unspecified |
| Mood disorder | F31.61 | Bipolar disorder, current episode mixed, mild |
| Mood disorder | F31.62 | Bipolar disorder, current episode mixed, moderate |
| Mood disorder | F31.63 | Bipolar disorder, current episode mixed, severe, without psychotic features |
| Mood and psychotic disorders | F31.64 | Bipolar disorder, current episode, mixed, severe with psychotic features |
| Mood disorder | F31.70 | Bipolar disorder, currently in remission, most recent episode unspecified |
| Mood disorder | F31.71 | Bipolar disorder, in partial remission, most recent episode hypomanic |
| Mood disorder | F31.72 | Bipolar disorder, in full remission, most recent episode hypomanic |
| Mood disorder | F31.73 | Bipolar disorder, in partial remission, most recent episode manic |
| Mood disorder | F31.74 | Bipolar disorder, in full remission, most recent episode manic |
| Mood disorder | F31.75 | Bipolar disorder, in partial remission, most recent episode depressed |
| Mood disorder | F31.76 | Bipolar disorder, in full remission, most recent episode depressed |
| Mood disorder | F31.77 | Bipolar disorder, in partial remission, most recent episode mixed |
| Mood disorder | F31.78 | Bipolar disorder, in full remission, most recent episode mixed |
| Mood disorder | F31.81 | Bipolar II disorder |
| Mood disorder | F31.89 | Other bipolar disorder |
| Mood disorder | F31.9 | Bipolar disorder, unspecified |
| Mood disorder | F32.0 | Major depressive disorder, single episode, mild |
| Mood disorder | F32.1 | Major depressive disorder, single episode, moderate |
| Mood disorder | F32.2 | Major depressive disorder, single episode, severe without psychotic features |
| Mood and psychotic disorders | F32.3 | Major depressive disorder, single episode, severe with psychotic features |
| Mood disorder | F32.4 | Major depressive disorder, single episode, in partial remission |
| Mood disorder | F32.5 | Major depressive disorder, single episode, in full remission |
| Mood disorder | F32.81 | Premenstrual dysphoric disorder |
| Mood disorder | F32.89 | Other specified depressive episodes |
| Mood disorder | F32.9 | Major depressive disorder, single episode, unspecified |
| Mood disorder | F32.A | Depression, unspecified |
| Mood disorder | F33.0 | Major depressive disorder, recurrent, mild |
| Mood disorder | F33.1 | Major depressive disorder, recurrent, moderate |
| Mood disorder | F33.2 | Major depressive disorder, recurrent, severe without psychotic features |
| Mood and psychotic disorders | F33.3 | Major depressive disorder, recurrent, severe with psychotic symptoms |
| Mood disorder | F33.40 | Major depressive disorder, recurrent, in remission, unspecified |
| Mood disorder | F33.41 | Major depressive disorder, recurrent, in partial remission |
| Mood disorder | F33.42 | Major depressive disorder, recurrent, in full remission |
| Mood disorder | F33.8 | Other recurrent depressive disorders |
| Mood disorder | F33.9 | Major depressive disorder, recurrent, unspecified |
| Mood disorder | F34.0 | Cyclothymic disorder |
| Mood disorder | F34.1 | Dysthymic disorder |
| Mood disorder | F34.81 | Disruptive mood dysregulation disorder |
| Mood disorder | F34.89 | Other specified persistent mood disorders |
| Mood disorder | F34.9 | Persistent mood disorder, unspecified |
| Mood disorder | F39 | Unspecified mood disorder |
| Mood disorder | F53.0 | Postpartum depression |
| Psychotic disorder | F53.1 | Puerperal psychosis |

*Note.* The diagnostic category indicates the study phenotype definition(s) to which each code was assigned. ICD-10-CM = International Classification of Diseases, Tenth Revision, Clinical Modification.

**SUPPLEMENTARY TABLE S2 | ICD-10-CM codes used to define the primary outcome of suicide attempt.**

All codes indicating intentional self-harm or suicide attempt were included; any patient with at least one recorded code was classified as having a suicide attempt for the purposes of this study.

| **ICD-10-CM Code** | **Description** |
| --- | --- |
| T36.0X2A | Poisoning by penicillins, intentional self-harm, initial encounter |
| T36.1X2A | Poisoning by cephalosporins and other beta-lactam antibiotics, intentional self-harm, initial encounter |
| T36.2X2A | Poisoning by chloramphenicol group, intentional self-harm, initial encounter |
| T36.3X2A | Poisoning by macrolides, intentional self-harm, initial encounter |
| T36.4X2A | Poisoning by tetracyclines, intentional self-harm, initial encounter |
| T36.5X2A | Poisoning by aminoglycosides, intentional self-harm, initial encounter |
| T36.6X2A | Poisoning by rifampicins, intentional self-harm, initial encounter |
| T36.7X2A | Poisoning by antifungal antibiotics, systemically used, intentional self-harm, initial encounter |
| T36.8X2A | Poisoning by other systemic antibiotics, intentional self-harm, initial encounter |
| T36.92XA | Poisoning by unspecified systemic antibiotic, intentional self-harm, initial encounter |
| T37.0X2A | Poisoning by sulfonamides, intentional self-harm, initial encounter |
| T37.1X2A | Poisoning by antimycobacterial drugs, intentional self-harm, initial encounter |
| T37.2X2A | Poisoning by antimalarials and drugs acting on other blood protozoa, intentional self-harm, initial encounter |
| T37.3X2A | Poisoning by other antiprotozoal drugs, intentional self-harm, initial encounter |
| T37.4X2A | Poisoning by anthelminthics, intentional self-harm, initial encounter |
| T37.5X2A | Poisoning by antiviral drugs, intentional self-harm, initial encounter |
| T37.8X2A | Poisoning by other specified systemic anti-infectives and antiparasitics, intentional self-harm, initial encounter |
| T37.92XA | Poisoning by unspecified systemic anti-infective and antiparasitics, intentional self-harm, initial encounter |
| T38.0X2A | Poisoning by glucocorticoids and synthetic analogues, intentional self-harm, initial encounter |
| T38.1X2A | Poisoning by thyroid hormones and substitutes, intentional self-harm, initial encounter |
| T38.2X2A | Poisoning by antithyroid drugs, intentional self-harm, initial encounter |
| T38.3X2A | Poisoning by insulin and oral hypoglycemic [antidiabetic] drugs, intentional self-harm, initial encounter |
| T38.4X2A | Poisoning by oral contraceptives, intentional self-harm, initial encounter |
| T38.5X2A | Poisoning by other estrogens and progestogens, intentional self-harm, initial encounter |
| T38.6X2A | Poisoning by antigonadotrophins, antiestrogens, antiandrogens, not elsewhere classified, intentional self-harm, initial encounter |
| T38.7X2A | Poisoning by androgens and anabolic congeners, intentional self-harm, initial encounter |
| T38.802A | Poisoning by unspecified hormones and synthetic substitutes, intentional self-harm, initial encounter |
| T38.812A | Poisoning by anterior pituitary [adenohypophyseal] hormones, intentional self-harm, initial encounter |
| T38.892A | Poisoning by other hormones and synthetic substitutes, intentional self-harm, initial encounter |
| T38.902A | Poisoning by unspecified hormone antagonists, intentional self-harm, initial encounter |
| T38.992A | Poisoning by other hormone antagonists, intentional self-harm, initial encounter |
| T39.012A | Poisoning by aspirin, intentional self-harm, initial encounter |
| T39.092A | Poisoning by salicylates, intentional self-harm, initial encounter |
| T39.1X2A | Poisoning by 4-Aminophenol derivatives, intentional self-harm, initial encounter |
| T39.2X2A | Poisoning by pyrazolone derivatives, intentional self-harm, initial encounter |
| T39.312A | Poisoning by propionic acid derivatives, intentional self-harm, initial encounter |
| T39.392A | Poisoning by other nonsteroidal anti-inflammatory drugs [NSAID], intentional self-harm, initial encounter |
| T39.4X2A | Poisoning by antirheumatics, not elsewhere classified, intentional self-harm, initial encounter |
| T39.8X2A | Poisoning by other nonopioid analgesics and antipyretics, not elsewhere classified, intentional self-harm, initial encounter |
| T39.92XA | Poisoning by unspecified nonopioid analgesic, antipyretic and antirheumatic, intentional self-harm, initial encounter |
| T40.0X2A | Poisoning by opium, intentional self-harm, initial encounter |
| T40.1X2A | Poisoning by heroin, intentional self-harm, initial encounter |
| T40.2X2A | Poisoning by other opioids, intentional self-harm, initial encounter |
| T40.3X2A | Poisoning by methadone, intentional self-harm, initial encounter |
| T40.412A | POISONING BY FENTANYL/ANALOG SELF-HARM INITIAL |
| T40.422A | POISONING BY TRAMADOL INTENTIONAL SELF-HARM INIT |
| T40.492A | POISONING OTH SYNTH NARCOTICS SELF-HARM INIT |
| T40.4X2A | Poisoning by other synthetic narcotics, intentional self-harm, initial encounter |
| T40.5X2A | Poisoning by cocaine, intentional self-harm, initial encounter |
| T40.602A | Poisoning by unspecified narcotics, intentional self-harm, initial encounter |
| T40.692A | Poisoning by other narcotics, intentional self-harm, initial encounter |
| T40.712A | POISONING BY CANNABIS INTNTNL SELF-HARM INIT |
| T40.722A | POISON/SYNTH CANNABINOIDS INTENT SLF-HRM INIT |
| T40.7X2A | Poisoning by cannabis (derivatives), intentional self-harm, initial encounter |
| T40.8X2A | Poisoning by lysergide [LSD], intentional self-harm, initial encounter |
| T40.902A | Poisoning by unspecified psychodysleptics [hallucinogens], intentional self-harm, initial encounter |
| T40.992A | Poisoning by other psychodysleptics [hallucinogens], intentional self-harm, initial encounter |
| T41.0X2A | Poisoning by inhaled anesthetics, intentional self-harm, initial encounter |
| T41.1X2A | Poisoning by intravenous anesthetics, intentional self-harm, initial encounter |
| T41.202A | Poisoning by unspecified general anesthetics, intentional self-harm, initial encounter |
| T41.292A | Poisoning by other general anesthetics, intentional self-harm, initial encounter |
| T41.3X2A | Poisoning by local anesthetics, intentional self-harm, initial encounter |
| T41.42XA | Poisoning by unspecified anesthetic, intentional self-harm, initial encounter |
| T41.5X2A | Poisoning by therapeutic gases, intentional self-harm, initial encounter |
| T42.0X2A | Poisoning by hydantoin derivatives, intentional self-harm, initial encounter |
| T42.1X2A | Poisoning by iminostilbenes, intentional self-harm, initial encounter |
| T42.2X2A | Poisoning by succinimides and oxazolidinediones, intentional self-harm, initial encounter |
| T42.3X2A | Poisoning by barbiturates, intentional self-harm, initial encounter |
| T42.4X2A | Poisoning by benzodiazepines, intentional self-harm, initial encounter |
| T42.5X2A | Poisoning by mixed antiepileptics, intentional self-harm, initial encounter |
| T42.6X2A | Poisoning by other antiepileptic and sedative-hypnotic drugs, intentional self-harm, initial encounter |
| T42.72XA | Poisoning by unspecified antiepileptic and sedative-hypnotic drugs, intentional self-harm, initial encounter |
| T42.8X2A | Poisoning by antiparkinsonism drugs and other central muscle-tone depressants, intentional self-harm, initial encounter |
| T43.012A | Poisoning by tricyclic antidepressants, intentional self-harm, initial encounter |
| T43.022A | Poisoning by tetracyclic antidepressants, intentional self-harm, initial encounter |
| T43.1X2A | Poisoning by monoamine-oxidase-inhibitor antidepressants, intentional self-harm, initial encounter |
| T43.202A | Poisoning by unspecified antidepressants, intentional self-harm, initial encounter |
| T43.212A | Poisoning by selective serotonin and norepinephrine reuptake inhibitors, intentional self-harm, initial encounter |
| T43.222A | Poisoning by selective serotonin reuptake inhibitors, intentional self-harm, initial encounter |
| T43.292A | Poisoning by other antidepressants, intentional self-harm, initial encounter |
| T43.3X2A | Poisoning by phenothiazine antipsychotics and neuroleptics, intentional self-harm, initial encounter |
| T43.4X2A | Poisoning by butyrophenone and thiothixene neuroleptics, intentional self-harm, initial encounter |
| T43.502A | Poisoning by unspecified antipsychotics and neuroleptics, intentional self-harm, initial encounter |
| T43.592A | Poisoning by other antipsychotics and neuroleptics, intentional self-harm, initial encounter |
| T43.602A | Poisoning by unspecified psychostimulants, intentional self-harm, initial encounter |
| T43.612A | Poisoning by caffeine, intentional self-harm, initial encounter |
| T43.622A | Poisoning by amphetamines, intentional self-harm, initial encounter |
| T43.632A | Poisoning by methylphenidate, intentional self-harm, initial encounter |
| T43.642A | POISONING BY ECSTASY SELF-HARM INITIAL |
| T43.652A | POISONING METHAMPHETAMINES SELF-HARM INITIAL ENC |
| T43.692A | Poisoning by other psychostimulants, intentional self-harm, initial encounter |
| T43.8X2A | Poisoning by other psychotropic drugs, intentional self-harm, initial encounter |
| T43.92XA | Poisoning by unspecified psychotropic drug, intentional self-harm, initial encounter |
| T44.0X2A | Poisoning by anticholinesterase agents, intentional self-harm, initial encounter |
| T44.1X2A | Poisoning by other parasympathomimetics [cholinergics], intentional self-harm, initial encounter |
| T44.2X2A | Poisoning by ganglionic blocking drugs, intentional self-harm, initial encounter |
| T44.3X2A | Poisoning by other parasympatholytics [anticholinergics and antimuscarinics] and spasmolytics, intentional self-harm, initial encounter |
| T44.4X2A | Poisoning by predominantly alpha-adrenoreceptor agonists, intentional self-harm, initial encounter |
| T44.5X2A | Poisoning by predominantly beta-adrenoreceptor agonists, intentional self-harm, initial encounter |
| T44.6X2A | Poisoning by alpha-adrenoreceptor antagonists, intentional self-harm, initial encounter |
| T44.7X2A | Poisoning by beta-adrenoreceptor antagonists, intentional self-harm, initial encounter |
| T44.8X2A | Poisoning by centrally-acting and adrenergic-neuron-blocking agents, intentional self-harm, initial encounter |
| T44.902A | Poisoning by unspecified drugs primarily affecting the autonomic nervous system, intentional self-harm, initial encounter |
| T44.992A | Poisoning by other drug primarily affecting the autonomic nervous system, intentional self-harm, initial encounter |
| T45.0X2A | Poisoning by antiallergic and antiemetic drugs, intentional self-harm, initial encounter |
| T45.1X2A | Poisoning by antineoplastic and immunosuppressive drugs, intentional self-harm, initial encounter |
| T45.2X2A | Poisoning by vitamins, intentional self-harm, initial encounter |
| T45.3X2A | Poisoning by enzymes, intentional self-harm, initial encounter |
| T45.4X2A | Poisoning by iron and its compounds, intentional self-harm, initial encounter |
| T45.512A | Poisoning by anticoagulants, intentional self-harm, initial encounter |
| T45.522A | Poisoning by antithrombotic drugs, intentional self-harm, initial encounter |
| T45.602A | Poisoning by unspecified fibrinolysis-affecting drugs, intentional self-harm, initial encounter |
| T45.612A | Poisoning by thrombolytic drug, intentional self-harm, initial encounter |
| T45.622A | Poisoning by hemostatic drug, intentional self-harm, initial encounter |
| T45.692A | Poisoning by other fibrinolysis-affecting drugs, intentional self-harm, initial encounter |
| T45.7X2A | Poisoning by anticoagulant antagonists, vitamin K and other coagulants, intentional self-harm, initial encounter |
| T45.8X2A | Poisoning by other primarily systemic and hematological agents, intentional self-harm, initial encounter |
| T45.92XA | Poisoning by unspecified primarily systemic and hematological agent, intentional self-harm, initial encounter |
| T46.0X2A | Poisoning by cardiac-stimulant glycosides and drugs of similar action, intentional self-harm, initial encounter |
| T46.1X2A | Poisoning by calcium-channel blockers, intentional self-harm, initial encounter |
| T46.2X2A | Poisoning by other antidysrhythmic drugs, intentional self-harm, initial encounter |
| T46.3X2A | Poisoning by coronary vasodilators, intentional self-harm, initial encounter |
| T46.4X2A | Poisoning by angiotensin-converting-enzyme inhibitors, intentional self-harm, initial encounter |
| T46.5X2A | Poisoning by other antihypertensive drugs, intentional self-harm, initial encounter |
| T46.6X2A | Poisoning by antihyperlipidemic and antiarteriosclerotic drugs, intentional self-harm, initial encounter |
| T46.7X2A | Poisoning by peripheral vasodilators, intentional self-harm, initial encounter |
| T46.8X2A | Poisoning by antivaricose drugs, including sclerosing agents, intentional self-harm, initial encounter |
| T46.902A | Poisoning by unspecified agents primarily affecting the cardiovascular system, intentional self-harm, initial encounter |
| T46.992A | Poisoning by other agents primarily affecting the cardiovascular system, intentional self-harm, initial encounter |
| T47.0X2A | Poisoning by histamine H2-receptor blockers, intentional self-harm, initial encounter |
| T47.1X2A | Poisoning by other antacids and anti-gastric-secretion drugs, intentional self-harm, initial encounter |
| T47.2X2A | Poisoning by stimulant laxatives, intentional self-harm, initial encounter |
| T47.3X2A | Poisoning by saline and osmotic laxatives, intentional self-harm, initial encounter |
| T47.4X2A | Poisoning by other laxatives, intentional self-harm, initial encounter |
| T47.5X2A | Poisoning by digestants, intentional self-harm, initial encounter |
| T47.6X2A | Poisoning by antidiarrheal drugs, intentional self-harm, initial encounter |
| T47.7X2A | Poisoning by emetics, intentional self-harm, initial encounter |
| T47.8X2A | Poisoning by other agents primarily affecting gastrointestinal system, intentional self-harm, initial encounter |
| T47.92XA | Poisoning by unspecified agents primarily affecting the gastrointestinal system, intentional self-harm, initial encounter |
| T48.0X2A | Poisoning by oxytocic drugs, intentional self-harm, initial encounter |
| T48.1X2A | Poisoning by skeletal muscle relaxants [neuromuscular blocking agents], intentional self-harm, initial encounter |
| T48.202A | Poisoning by unspecified drugs acting on muscles, intentional self-harm, initial encounter |
| T48.292A | Poisoning by other drugs acting on muscles, intentional self-harm, initial encounter |
| T48.3X2A | Poisoning by antitussives, intentional self-harm, initial encounter |
| T48.4X2A | Poisoning by expectorants, intentional self-harm, initial encounter |
| T48.5X2A | Poisoning by other anti-common-cold drugs, intentional self-harm, initial encounter |
| T48.6X2A | Poisoning by antiasthmatics, intentional self-harm, initial encounter |
| T48.902A | Poisoning by unspecified agents primarily acting on the respiratory system, intentional self-harm, initial encounter |
| T48.992A | Poisoning by other agents primarily acting on the respiratory system, intentional self-harm, initial encounter |
| T49.0X2A | Poisoning by local antifungal, anti-infective and anti-inflammatory drugs, intentional self-harm, initial encounter |
| T49.1X2A | Poisoning by antipruritics, intentional self-harm, initial encounter |
| T49.2X2A | Poisoning by local astringents and local detergents, intentional self-harm, initial encounter |
| T49.3X2A | Poisoning by emollients, demulcents and protectants, intentional self-harm, initial encounter |
| T49.4X2A | Poisoning by keratolytics, keratoplastics, and other hair treatment drugs and preparations, intentional self-harm, initial encounter |
| T49.5X2A | Poisoning by ophthalmological drugs and preparations, intentional self-harm, initial encounter |
| T49.6X2A | Poisoning by otorhinolaryngological drugs and preparations, intentional self-harm, initial encounter |
| T49.7X2A | Poisoning by dental drugs, topically applied, intentional self-harm, initial encounter |
| T49.8X2A | Poisoning by other topical agents, intentional self-harm, initial encounter |
| T49.92XA | Poisoning by unspecified topical agent, intentional self-harm, initial encounter |
| T50.0X2A | Poisoning by mineralocorticoids and their antagonists, intentional self-harm, initial encounter |
| T50.1X2A | Poisoning by loop [high-ceiling] diuretics, intentional self-harm, initial encounter |
| T50.2X2A | Poisoning by carbonic-anhydrase inhibitors, benzothiadiazides and other diuretics, intentional self-harm, initial encounter |
| T50.3X2A | Poisoning by electrolytic, caloric and water-balance agents, intentional self-harm, initial encounter |
| T50.4X2A | Poisoning by drugs affecting uric acid metabolism, intentional self-harm, initial encounter |
| T50.5X2A | Poisoning by appetite depressants, intentional self-harm, initial encounter |
| T50.6X2A | Poisoning by antidotes and chelating agents, intentional self-harm, initial encounter |
| T50.7X2A | Poisoning by analeptics and opioid receptor antagonists, intentional self-harm, initial encounter |
| T50.8X2A | Poisoning by diagnostic agents, intentional self-harm, initial encounter |
| T50.902A | Poisoning by unspecified drugs, medicaments and biological substances, intentional self-harm, initial encounter |
| T50.912A | POISON MULT UNS DRUG/MEDS/BIO SBST SLF-HRM INIT |
| T50.992A | Poisoning by other drugs, medicaments and biological substances, intentional self-harm, initial encounter |
| T50.A12A | Poisoning by pertussis vaccine, including combinations with a pertussis component, intentional self-harm, initial encounter |
| T50.A22A | Poisoning by mixed bacterial vaccines without a pertussis component, intentional self-harm, initial encounter |
| T50.A92A | Poisoning by other bacterial vaccines, intentional self-harm, initial encounter |
| T50.B12A | Poisoning by smallpox vaccines, intentional self-harm, initial encounter |
| T50.B92A | Poisoning by other viral vaccines, intentional self-harm, initial encounter |
| T50.Z12A | Poisoning by immunoglobulin, intentional self-harm, initial encounter |
| T50.Z92A | Poisoning by other vaccines and biological substances, intentional self-harm, initial encounter |
| T51.0X2A | Toxic effect of ethanol, intentional self-harm, initial encounter |
| T51.1X2A | Toxic effect of methanol, intentional self-harm, initial encounter |
| T51.2X2A | Toxic effect of 2-Propanol, intentional self-harm, initial encounter |
| T51.3X2A | Toxic effect of fuel oil, intentional self-harm, initial encounter |
| T51.8X2A | Toxic effect of other alcohols, intentional self-harm, initial encounter |
| T51.92XA | Toxic effect of unspecified alcohol, intentional self-harm, initial encounter |
| T52.0X2A | Toxic effect of petroleum products, intentional self-harm, initial encounter |
| T52.1X2A | Toxic effect of benzene, intentional self-harm, initial encounter |
| T52.2X2A | Toxic effect of homologues of benzene, intentional self-harm, initial encounter |
| T52.3X2A | Toxic effect of glycols, intentional self-harm, initial encounter |
| T52.4X2A | Toxic effect of ketones, intentional self-harm, initial encounter |
| T52.8X2A | Toxic effect of other organic solvents, intentional self-harm, initial encounter |
| T52.92XA | Toxic effect of unspecified organic solvent, intentional self-harm, initial encounter |
| T53.0X2A | Toxic effect of carbon tetrachloride, intentional self-harm, initial encounter |
| T53.1X2A | Toxic effect of chloroform, intentional self-harm, initial encounter |
| T53.2X2A | Toxic effect of trichloroethylene, intentional self-harm, initial encounter |
| T53.3X2A | Toxic effect of tetrachloroethylene, intentional self-harm, initial encounter |
| T53.4X2A | Toxic effect of dichloromethane, intentional self-harm, initial encounter |
| T53.5X2A | Toxic effect of chlorofluorocarbons, intentional self-harm, initial encounter |
| T53.6X2A | Toxic effect of other halogen derivatives of aliphatic hydrocarbons, intentional self-harm, initial encounter |
| T53.7X2A | Toxic effect of other halogen derivatives of aromatic hydrocarbons, intentional self-harm, initial encounter |
| T53.92XA | Toxic effect of unspecified halogen derivatives of aliphatic and aromatic hydrocarbons, intentional self-harm, initial encounter |
| T54.0X2A | Toxic effect of phenol and phenol homologues, intentional self-harm, initial encounter |
| T54.1X2A | Toxic effect of other corrosive organic compounds, intentional self-harm, initial encounter |
| T54.2X2A | Toxic effect of corrosive acids and acid-like substances, intentional self-harm, initial encounter |
| T54.3X2A | Toxic effect of corrosive alkalis and alkali-like substances, intentional self-harm, initial encounter |
| T54.92XA | Toxic effect of unspecified corrosive substance, intentional self-harm, initial encounter |
| T55.0X2A | Toxic effect of soaps, intentional self-harm, initial encounter |
| T55.1X2A | Toxic effect of detergents, intentional self-harm, initial encounter |
| T56.0X2A | Toxic effect of lead and its compounds, intentional self-harm, initial encounter |
| T56.1X2A | Toxic effect of mercury and its compounds, intentional self-harm, initial encounter |
| T56.2X2A | Toxic effect of chromium and its compounds, intentional self-harm, initial encounter |
| T56.3X2A | Toxic effect of cadmium and its compounds, intentional self-harm, initial encounter |
| T56.4X2A | Toxic effect of copper and its compounds, intentional self-harm, initial encounter |
| T56.5X2A | Toxic effect of zinc and its compounds, intentional self-harm, initial encounter |
| T56.6X2A | Toxic effect of tin and its compounds, intentional self-harm, initial encounter |
| T56.7X2A | Toxic effect of beryllium and its compounds, intentional self-harm, initial encounter |
| T56.812A | Toxic effect of thallium, intentional self-harm, initial encounter |
| T56.822A | TOX EFFECT GADOLINIUM INTENT SELF-HARM INT ENC |
| T56.892A | Toxic effect of other metals, intentional self-harm, initial encounter |
| T56.92XA | Toxic effect of unspecified metal, intentional self-harm, initial encounter |
| T57.0X2A | Toxic effect of arsenic and its compounds, intentional self-harm, initial encounter |
| T57.1X2A | Toxic effect of phosphorus and its compounds, intentional self-harm, initial encounter |
| T57.2X2A | Toxic effect of manganese and its compounds, intentional self-harm, initial encounter |
| T57.3X2A | Toxic effect of hydrogen cyanide, intentional self-harm, initial encounter |
| T57.8X2A | Toxic effect of other specified inorganic substances, intentional self-harm, initial encounter |
| T57.92XA | Toxic effect of unspecified inorganic substance, intentional self-harm, initial encounter |
| T58.02XA | Toxic effect of carbon monoxide from motor vehicle exhaust, intentional self-harm, initial encounter |
| T58.12XA | Toxic effect of carbon monoxide from utility gas, intentional self-harm, initial encounter |
| T58.2X2A | Toxic effect of carbon monoxide from incomplete combustion of other domestic fuels, intentional self-harm, initial encounter |
| T58.8X2A | Toxic effect of carbon monoxide from other source, intentional self-harm, initial encounter |
| T58.92XA | Toxic effect of carbon monoxide from unspecified source, intentional self-harm, initial encounter |
| T59.0X2A | Toxic effect of nitrogen oxides, intentional self-harm, initial encounter |
| T59.1X2A | Toxic effect of sulfur dioxide, intentional self-harm, initial encounter |
| T59.2X2A | Toxic effect of formaldehyde, intentional self-harm, initial encounter |
| T59.3X2A | Toxic effect of lacrimogenic gas, intentional self-harm, initial encounter |
| T59.4X2A | Toxic effect of chlorine gas, intentional self-harm, initial encounter |
| T59.5X2A | Toxic effect of fluorine gas and hydrogen fluoride, intentional self-harm, initial encounter |
| T59.6X2A | Toxic effect of hydrogen sulfide, intentional self-harm, initial encounter |
| T59.7X2A | Toxic effect of carbon dioxide, intentional self-harm, initial encounter |
| T59.812A | Toxic effect of smoke, intentional self-harm, initial encounter |
| T59.892A | Toxic effect of other specified gases, fumes and vapors, intentional self-harm, initial encounter |
| T59.92XA | Toxic effect of unspecified gases, fumes and vapors, intentional self-harm, initial encounter |
| T60.0X2A | Toxic effect of organophosphate and carbamate insecticides, intentional self-harm, initial encounter |
| T60.1X2A | Toxic effect of halogenated insecticides, intentional self-harm, initial encounter |
| T60.2X2A | Toxic effect of other insecticides, intentional self-harm, initial encounter |
| T60.3X2A | Toxic effect of herbicides and fungicides, intentional self-harm, initial encounter |
| T60.4X2A | Toxic effect of rodenticides, intentional self-harm, initial encounter |
| T60.8X2A | Toxic effect of other pesticides, intentional self-harm, initial encounter |
| T60.92XA | Toxic effect of unspecified pesticide, intentional self-harm, initial encounter |
| T61.02XA | Ciguatera fish poisoning, intentional self-harm, initial encounter |
| T61.12XA | Scombroid fish poisoning, intentional self-harm, initial encounter |
| T61.772A | Other fish poisoning, intentional self-harm, initial encounter |
| T61.782A | Other shellfish poisoning, intentional self-harm, initial encounter |
| T61.8X2A | Toxic effect of other seafood, intentional self-harm, initial encounter |
| T61.92XA | Toxic effect of unspecified seafood, intentional self-harm, initial encounter |
| T62.0X2A | Toxic effect of ingested mushrooms, intentional self-harm, initial encounter |
| T62.1X2A | Toxic effect of ingested berries, intentional self-harm, initial encounter |
| T62.2X2A | Toxic effect of other ingested (parts of) plant(s), intentional self-harm, initial encounter |
| T62.8X2A | Toxic effect of other specified noxious substances eaten as food, intentional self-harm, initial encounter |
| T62.92XA | Toxic effect of unspecified noxious substance eaten as food, intentional self-harm, initial encounter |
| T63.002A | Toxic effect of unspecified snake venom, intentional self-harm, initial encounter |
| T63.012A | Toxic effect of rattlesnake venom, intentional self-harm, initial encounter |
| T63.022A | Toxic effect of coral snake venom, intentional self-harm, initial encounter |
| T63.032A | Toxic effect of taipan venom, intentional self-harm, initial encounter |
| T63.042A | Toxic effect of cobra venom, intentional self-harm, initial encounter |
| T63.062A | Toxic effect of venom of other North and South American snake, intentional self-harm, initial encounter |
| T63.072A | Toxic effect of venom of other Australian snake, intentional self-harm, initial encounter |
| T63.082A | Toxic effect of venom of other African and Asian snake, intentional self-harm, initial encounter |
| T63.092A | Toxic effect of venom of other snake, intentional self-harm, initial encounter |
| T63.112A | Toxic effect of venom of Gila monster, intentional self-harm, initial encounter |
| T63.122A | Toxic effect of venom of other venomous lizard, intentional self-harm, initial encounter |
| T63.192A | Toxic effect of venom of other reptiles, intentional self-harm, initial encounter |
| T63.2X2A | Toxic effect of venom of scorpion, intentional self-harm, initial encounter |
| T63.302A | Toxic effect of unspecified spider venom, intentional self-harm, initial encounter |
| T63.312A | Toxic effect of venom of black widow spider, intentional self-harm, initial encounter |
| T63.322A | Toxic effect of venom of tarantula, intentional self-harm, initial encounter |
| T63.332A | Toxic effect of venom of brown recluse spider, intentional self-harm, initial encounter |
| T63.392A | Toxic effect of venom of other spider, intentional self-harm, initial encounter |
| T63.412A | Toxic effect of venom of centipedes and venomous millipedes, intentional self-harm, initial encounter |
| T63.422A | Toxic effect of venom of ants, intentional self-harm, initial encounter |
| T63.432A | Toxic effect of venom of caterpillars, intentional self-harm, initial encounter |
| T63.442A | Toxic effect of venom of bees, intentional self-harm, initial encounter |
| T63.452A | Toxic effect of venom of hornets, intentional self-harm, initial encounter |
| T63.462A | Toxic effect of venom of wasps, intentional self-harm, initial encounter |
| T63.482A | Toxic effect of venom of other arthropod, intentional self-harm, initial encounter |
| T63.512A | Toxic effect of contact with stingray, intentional self-harm, initial encounter |
| T63.592A | Toxic effect of contact with other venomous fish, intentional self-harm, initial encounter |
| T63.612A | Toxic effect of contact with Portugese Man-o-war, intentional self-harm, initial encounter |
| T63.622A | Toxic effect of contact with other jellyfish, intentional self-harm, initial encounter |
| T63.632A | Toxic effect of contact with sea anemone, intentional self-harm, initial encounter |
| T63.692A | Toxic effect of contact with other venomous marine animals, intentional self-harm, initial encounter |
| T63.712A | Toxic effect of contact with venomous marine plant, intentional self-harm, initial encounter |
| T63.792A | Toxic effect of contact with other venomous plant, intentional self-harm, initial encounter |
| T63.812A | Toxic effect of contact with venomous frog, intentional self-harm, initial encounter |
| T63.822A | Toxic effect of contact with venomous toad, intentional self-harm, initial encounter |
| T63.832A | Toxic effect of contact with other venomous amphibian, intentional self-harm, initial encounter |
| T63.892A | Toxic effect of contact with other venomous animals, intentional self-harm, initial encounter |
| T63.92XA | Toxic effect of contact with unspecified venomous animal, intentional self-harm, initial encounter |
| T64.02XA | Toxic effect of aflatoxin, intentional self-harm, initial encounter |
| T64.82XA | Toxic effect of other mycotoxin food contaminants, intentional self-harm, initial encounter |
| T65.0X2A | Toxic effect of cyanides, intentional self-harm, initial encounter |
| T65.1X2A | Toxic effect of strychnine and its salts, intentional self-harm, initial encounter |
| T65.212A | Toxic effect of chewing tobacco, intentional self-harm, initial encounter |
| T65.222A | Toxic effect of tobacco cigarettes, intentional self-harm, initial encounter |
| T65.292A | Toxic effect of other tobacco and nicotine, intentional self-harm, initial encounter |
| T65.3X2A | Toxic effect of nitro derivatives and amino derivatives of benzene and its homologues, intentional self-harm, initial encounter |
| T65.4X2A | Toxic effect of carbon disulfide, intentional self-harm, initial encounter |
| T65.5X2A | Toxic effect of nitroglycerin and other nitric acids and esters, intentional self-harm, initial encounter |
| T65.6X2A | Toxic effect of paints and dyes, not elsewhere classified, intentional self-harm, initial encounter |
| T65.812A | Toxic effect of latex, intentional self-harm, initial encounter |
| T65.822A | Toxic effect of harmful algae and algae toxins, intentional self-harm, initial encounter |
| T65.832A | Toxic effect of fiberglass, intentional self-harm, initial encounter |
| T65.892A | Toxic effect of other specified substances, intentional self-harm, initial encounter |
| T65.92XA | Toxic effect of unspecified substance, intentional self-harm, initial encounter |
| T71.112A | Asphyxiation due to smothering under pillow, intentional self-harm, initial encounter |
| T71.122A | Asphyxiation due to plastic bag, intentional self-harm, initial encounter |
| T71.132A | Asphyxiation due to being trapped in bed linens, intentional self-harm, initial encounter |
| T71.152A | Asphyxiation due to smothering in furniture, intentional self-harm, initial encounter |
| T71.162A | Asphyxiation due to hanging, intentional self-harm, initial encounter |
| T71.192A | Asphyxiation due to mechanical threat to breathing due to other causes, intentional self-harm, initial encounter |
| T71.222A | Asphyxiation due to being trapped in a car trunk, intentional self-harm, initial encounter |
| T71.232A | Asphyxiation due to being trapped in a (discarded) refrigerator, intentional self-harm, initial encounter |
| X71.0XXA | Intentional self-harm by drowning and submersion while in bathtub, initial encounter |
| X71.1XXA | Intentional self-harm by drowning and submersion while in swimming pool, initial encounter |
| X71.2XXA | Intentional self-harm by drowning and submersion after jump into swimming pool, initial encounter |
| X71.3XXA | Intentional self-harm by drowning and submersion in natural water, initial encounter |
| X71.8XXA | Other intentional self-harm by drowning and submersion, initial encounter |
| X71.9XXA | Intentional self-harm by drowning and submersion, unspecified, initial encounter |
| X72.XXXA | Intentional self-harm by handgun discharge, initial encounter |
| X73.0XXA | Intentional self-harm by shotgun discharge, initial encounter |
| X73.1XXA | Intentional self-harm by hunting rifle discharge, initial encounter |
| X73.2XXA | Intentional self-harm by machine gun discharge, initial encounter |
| X73.8XXA | Intentional self-harm by other larger firearm discharge, initial encounter |
| X73.9XXA | Intentional self-harm by unspecified larger firearm discharge, initial encounter |
| X74.01XA | Intentional self-harm by air gun, initial encounter |
| X74.02XA | Intentional self-harm by paintball gun, initial encounter |
| X74.09XA | Intentional self-harm by other gas, air or spring-operated gun, initial encounter |
| X74.8XXA | Intentional self-harm by other firearm discharge, initial encounter |
| X74.9XXA | Intentional self-harm by unspecified firearm discharge, initial encounter |
| X75.XXXA | Intentional self-harm by explosive material, initial encounter |
| X76.XXXA | Intentional self-harm by smoke, fire and flames, initial encounter |
| X77.0XXA | Intentional self-harm by steam or hot vapors, initial encounter |
| X77.1XXA | Intentional self-harm by hot tap water, initial encounter |
| X77.2XXA | Intentional self-harm by other hot fluids, initial encounter |
| X77.3XXA | Intentional self-harm by hot household appliances, initial encounter |
| X77.8XXA | Intentional self-harm by other hot objects, initial encounter |
| X77.9XXA | Intentional self-harm by unspecified hot objects, initial encounter |
| X78.0XXA | Intentional self-harm by sharp glass, initial encounter |
| X78.1XXA | Intentional self-harm by knife, initial encounter |
| X78.2XXA | Intentional self-harm by sword or dagger, initial encounter |
| X78.8XXA | Intentional self-harm by other sharp object, initial encounter |
| X78.9XXA | Intentional self-harm by unspecified sharp object, initial encounter |
| X79.XXXA | Intentional self-harm by blunt object, initial encounter |
| X80.XXXA | Intentional self-harm by jumping from a high place, initial encounter |
| X81.0XXA | Intentional self-harm by jumping or lying in front of motor vehicle, initial encounter |
| X81.1XXA | Intentional self-harm by jumping or lying in front of (subway) train, initial encounter |
| X81.8XXA | Intentional self-harm by jumping or lying in front of other moving object, initial encounter |
| X82.0XXA | Intentional collision of motor vehicle with other motor vehicle, initial |
| X82.1XXA | Intentional collision of motor vehicle with train, initial encounter |
| X82.2XXA | Intentional collision of motor vehicle with tree, initial encounter |
| X82.8XXA | Other intentional self-harm by crashing of motor vehicle, initial encounter |
| X83.0XXA | Intentional self-harm by crashing of aircraft, initial encounter |
| X83.1XXA | Intentional self-harm by electrocution, initial encounter |
| X83.2XXA | Intentional self-harm by exposure to extremes of cold, initial encounter |
| X83.8XXA | Intentional self-harm by other specified means, initial encounter |
| T14.91 | Suicide attempt |
| T14.91XA | Suicide attempt initial encounter |

Note. ICD-10-CM = International Classification of Diseases, Tenth Revision, Clinical Modification. Code descriptions reflect the CDC's surveillance case definition for nonfatal suicide attempts (Hedegaard et al., 2018).

**SUPPLEMENTARY TABLE S3 | Data dictionary of candidate predictor variables.**

Provided as a separate file (Supplementary Table 3.xlsx) due to file size. Lists all patient-level and encounter-level variables considered as candidate predictors prior to feature selection and ablation.

**SUPPLEMENTARY TABLE S4. Imputation sensitivity analysis.**

AUROC, AUPRC, Recall@99Spec, Precision@top1%, Calibration Slope, Balanced Accuracy, and Standardized Net Benefit for each algorithm under Zero, Mean, and Median imputation (pre-ablation feature set, CSL=False, Isotonic calibration). Balanced Accuracy and Standardized Net Benefit are evaluated at the policy decision threshold (p_t = 1/11), consistent with the net-benefit weighting used throughout.

| **Algorithm** | **Impute** | **AUROC** | **AUPRC** | **Recall@99Spec** | **Precision@top1%** | **Calibration Slope** | **Balanced Accuracy** | **Std. Net Benefit** |
| --- | --- | --- | --- | --- | --- | --- | --- | --- |
| ANN | Zero | 0.8707 | 0.1221 | 0.2513 | 0.2157 | 0.9675 | 0.6832 | 0.1760 |
| ANN | Mean | 0.8435 | 0.1055 | 0.1824 | 0.1950 | 0.9806 | 0.6220 | 0.1286 |
| ANN | Median | 0.8606 | 0.1101 | 0.2131 | 0.1961 | 0.9658 | 0.6240 | 0.1397 |
| EN | Zero | 0.9013 | 0.2073 | 0.4920 | 0.3478 | 0.9626 | 0.7411 | 0.3928 |
| EN | Mean | 0.9009 | 0.2180 | 0.4910 | 0.3457 | 0.9639 | 0.7406 | 0.3912 |
| EN | Median | 0.9011 | 0.2162 | 0.4910 | 0.3478 | 0.9645 | 0.7392 | 0.3914 |
| LogReg | Zero | 0.9051 | 0.2291 | 0.4592 | 0.3602 | 0.9674 | 0.7374 | 0.3759 |
| LogReg | Mean | 0.9051 | 0.2292 | 0.4592 | 0.3602 | 0.9674 | 0.7374 | 0.3759 |
| LogReg | Median | 0.9051 | 0.2292 | 0.4592 | 0.3602 | 0.9674 | 0.7374 | 0.3758 |
| SVM | Zero | 0.9008 | 0.2249 | 0.4857 | 0.3540 | 1.0022 | 0.7465 | 0.3859 |
| SVM | Mean | 0.9005 | 0.2247 | 0.4857 | 0.3540 | 1.0024 | 0.7465 | 0.3858 |
| SVM | Median | 0.9008 | 0.2238 | 0.4857 | 0.3540 | 1.0023 | 0.7465 | 0.3858 |
| XGB | Zero | 0.9116 | 0.2521 | 0.4825 | 0.3715 | 0.9999 | 0.7429 | 0.3897 |
| XGB | Mean | 0.9095 | 0.2606 | 0.4942 | 0.3715 | 0.9934 | 0.7467 | 0.3989 |
| XGB | Median | 0.9117 | 0.2603 | 0.4899 | 0.3643 | 0.9956 | 0.7400 | 0.3894 |

**SUPPLEMENTARY TABLE S5a. Threshold-invariant performance metrics.** AUROC, AUPRC, Calibration Slope, Calibration Intercept, expected calibration error (ECE), Brier score, Balanced Accuracy, and Standardized Net Benefit for each algorithm (Zero imputation, pre-ablation feature set, CSL=False, Isotonic calibration). Balanced Accuracy and Standardized Net Benefit are evaluated at the policy decision threshold (p_t = 1/11), consistent with the net-benefit weighting used throughout.

| **Algorithm** | **AUROC** | **AUPRC** | **Calibration Slope** | **Calibration Intercept** | **ECE** | **Brier Score** | **Balanced Accuracy** | **Std. Net Benefit** |
| --- | --- | --- | --- | --- | --- | --- | --- | --- |
| ANN | 0.8707 | 0.1221 | 0.9675 | -0.0415 | 0.0007 | 0.0090 | 0.6832 | 0.1760 |
| EN | 0.9013 | 0.2073 | 0.9626 | -0.0210 | 0.0009 | 0.0081 | 0.7411 | 0.3928 |
| LogReg | 0.9051 | 0.2291 | 0.9674 | -0.0392 | 0.0006 | 0.0081 | 0.7374 | 0.3759 |
| SVM | 0.9008 | 0.2249 | 1.0022 | -0.0301 | 0.0005 | 0.0081 | 0.7465 | 0.3859 |
| XGB | 0.9116 | 0.2521 | 0.9999 | -0.0273 | 0.0004 | 0.0080 | 0.7429 | 0.3897 |

**SUPPLEMENTARY TABLE S5b. Threshold-dependent performance metrics.** Positive predictive value (PPV), sensitivity, specificity, Youden's index, and number needed to evaluate (NNE, the reciprocal of PPV) for each algorithm at seven prespecified operating points: the default probability threshold of 0.50; thresholds corresponding to the top 1%, 2%, and 5% of predicted risk; and thresholds corresponding to 99th-, 95th-, and 90th-percentile specificity.

| **Algorithm** | **Operating Point** | **PPV** | **Sensitivity** | **Specificity** | **Youden's Index** | **NNE** |
| --- | --- | --- | --- | --- | --- | --- |
| ANN | @0.50 (default) | 0.0500 | 0.0011 | 0.9998 | 0.0009 | 20.0 |
| ANN | @Precision1% | 0.2157 | 0.2216 | 0.9921 | 0.2137 | 4.64 |
| ANN | @Precision2% | 0.1668 | 0.3425 | 0.9832 | 0.3257 | 6.0 |
| ANN | @Precision5% | 0.0991 | 0.5090 | 0.9545 | 0.4635 | 10.09 |
| ANN | @99Spec | 0.2147 | 0.2513 | 0.9910 | 0.2423 | 4.66 |
| ANN | @95Spec | 0.0935 | 0.5217 | 0.9503 | 0.4720 | 10.7 |
| ANN | @90Spec | 0.0695 | 0.6119 | 0.9195 | 0.5314 | 14.39 |
| EN | @0.50 (default) | 0.1579 | 0.0032 | 0.9998 | 0.0030 | 6.33 |
| EN | @Precision1% | 0.3478 | 0.3574 | 0.9934 | 0.3508 | 2.88 |
| EN | @Precision2% | 0.2586 | 0.5313 | 0.9850 | 0.5163 | 3.87 |
| EN | @Precision5% | 0.1235 | 0.6341 | 0.9557 | 0.5898 | 8.1 |
| EN | @99Spec | 0.3312 | 0.4920 | 0.9902 | 0.4822 | 3.02 |
| EN | @95Spec | 0.1185 | 0.6479 | 0.9526 | 0.6005 | 8.44 |
| EN | @90Spec | 0.0785 | 0.7285 | 0.9159 | 0.6444 | 12.74 |
| LogReg | @0.50 (default) | 0.3696 | 0.0541 | 0.9991 | 0.0532 | 2.71 |
| LogReg | @Precision1% | 0.3602 | 0.3701 | 0.9935 | 0.3636 | 2.78 |
| LogReg | @Precision2% | 0.2561 | 0.5260 | 0.9850 | 0.5110 | 3.9 |
| LogReg | @Precision5% | 0.1249 | 0.6416 | 0.9558 | 0.5974 | 8.01 |
| LogReg | @99Spec | 0.3346 | 0.4592 | 0.9910 | 0.4502 | 2.99 |
| LogReg | @95Spec | 0.1852 | 0.5875 | 0.9746 | 0.5621 | 5.4 |
| LogReg | @90Spec | 0.0709 | 0.7317 | 0.9057 | 0.6374 | 14.1 |
| SVM | @0.50 (default) | 0.3571 | 0.0053 | 0.9999 | 0.0052 | 2.8 |
| SVM | @Precision1% | 0.3540 | 0.3637 | 0.9935 | 0.3572 | 2.82 |
| SVM | @Precision2% | 0.2550 | 0.5239 | 0.9850 | 0.5089 | 3.92 |
| SVM | @Precision5% | 0.1239 | 0.6363 | 0.9558 | 0.5921 | 8.07 |
| SVM | @99Spec | 0.3393 | 0.4857 | 0.9907 | 0.4764 | 2.95 |
| SVM | @95Spec | 0.1342 | 0.6331 | 0.9598 | 0.5929 | 7.45 |
| SVM | @90Spec | 0.0688 | 0.7381 | 0.9018 | 0.6399 | 14.53 |
| XGB | @0.50 (default) | 0.5000 | 0.0159 | 0.9998 | 0.0157 | 2.0 |
| XGB | @Precision1% | 0.3715 | 0.3818 | 0.9937 | 0.3755 | 2.69 |
| XGB | @Precision2% | 0.2633 | 0.5408 | 0.9851 | 0.5259 | 3.8 |
| XGB | @Precision5% | 0.1278 | 0.6564 | 0.9560 | 0.6124 | 7.82 |
| XGB | @99Spec | 0.3343 | 0.4825 | 0.9906 | 0.4731 | 2.99 |
| XGB | @95Spec | 0.1483 | 0.6373 | 0.9640 | 0.6013 | 6.74 |
| XGB | @90Spec | 0.0829 | 0.7285 | 0.9208 | 0.6493 | 12.06 |

**SUPPLEMENTARY TABLE S6 | Cost-sensitive learning (CSL) sensitivity analysis.**

AUROC, AUPRC, Recall@99Spec, Precision@top1%, Calibration Slope, Balanced Accuracy, and Standardized Net Benefit for each algorithm under CSL=False (baseline, matching Table 2a) and CSL=True (shaded rows), evaluated at the policy decision threshold (p_t = 1/11). Zero imputation, pre-ablation feature set, and Isotonic calibration throughout.

| **Algorithm** | **CSL** | **AUROC** | **AUPRC** | **Recall@99Spec** | **Precision@top1%** | **Calibration Slope** | **Balanced Accuracy** | **Std. Net Benefit** |
| --- | --- | --- | --- | --- | --- | --- | --- | --- |
| ANN | False | 0.8707 | 0.1221 | 0.2513 | 0.2157 | 0.9675 | 0.6832 | 0.1760 |
| ANN | True | 0.6975 | 0.0351 | 0.0329 | 0.0320 | 0.9578 | 0.5000 | 0.0000 |
| EN | False | 0.9013 | 0.2073 | 0.4920 | 0.3478 | 0.9626 | 0.7411 | 0.3928 |
| EN | True | 0.9015 | 0.2066 | 0.4899 | 0.3488 | 0.9613 | 0.7403 | 0.3950 |
| LogReg | False | 0.9051 | 0.2291 | 0.4592 | 0.3602 | 0.9674 | 0.7374 | 0.3759 |
| LogReg | True | 0.9009 | 0.2276 | 0.4581 | 0.3571 | 0.9621 | 0.7471 | 0.3783 |
| SVM | False | 0.9008 | 0.2249 | 0.4857 | 0.3540 | 1.0022 | 0.7465 | 0.3859 |
| SVM | True | 0.8958 | 0.2320 | 0.4857 | 0.3591 | 1.0004 | 0.7446 | 0.3854 |
| XGB | False | 0.9116 | 0.2521 | 0.4825 | 0.3715 | 0.9999 | 0.7429 | 0.3897 |
| XGB | True | 0.7399 | 0.2737 | 0.4889 | 0.3498 | 0.9949 | 0.7377 | 0.3917 |

**SUPPLEMENTARY TABLE S7 | Alternative calibration methods sensitivity analysis.**

AUROC, AUPRC, Recall@99Spec, Precision@top1%, Calibration Slope, Balanced Accuracy, and Standardized Net Benefit for each algorithm under all four calibration methods evaluated (Raw, Isotonic, Platt, Temperature; Isotonic was used as the primary method throughout, per Table 2). Zero imputation, pre-ablation feature set, and CSL=False throughout. Balanced Accuracy and Standardized Net Benefit are evaluated at the policy decision threshold (p_t = 1/11). SVM’s Raw (uncalibrated) probabilities are a naive sigmoid transform of the model’s decision function rather than a fitted probability estimate; the resulting poor calibration (Calibration Slope = 7.16) produces degenerate downstream threshold-based metrics (Balanced Accuracy, Standardized Net Benefit) for this row specifically.

| **Algorithm** | **Calibration Method** | **AUROC** | **AUPRC** | **Recall@99Spec** | **Precision@top1%** | **Calibration Slope** | **Balanced Accuracy** | **Std. Net Benefit** |
| --- | --- | --- | --- | --- | --- | --- | --- | --- |
| ANN | Raw | 0.8731 | 0.1262 | 0.2715 | 0.2147 | 2.1529 | 0.5176 | 0.0264 |
| ANN | Isotonic | 0.8707 | 0.1221 | 0.2513 | 0.2157 | 0.9675 | 0.6832 | 0.1760 |
| ANN | Platt | 0.8731 | 0.1262 | 0.2715 | 0.2147 | 1.0109 | 0.6423 | 0.1720 |
| ANN | Temperature | 0.8731 | 0.1262 | 0.2715 | 0.2147 | 2.0370 | 0.5108 | 0.0162 |
| EN | Raw | 0.9021 | 0.2225 | 0.4952 | 0.3488 | 1.1596 | 0.7326 | 0.3844 |
| EN | Isotonic | 0.9013 | 0.2073 | 0.4920 | 0.3478 | 0.9626 | 0.7411 | 0.3928 |
| EN | Platt | 0.9021 | 0.2225 | 0.4952 | 0.3488 | 0.9964 | 0.7388 | 0.3947 |
| EN | Temperature | 0.9021 | 0.2225 | 0.4952 | 0.3488 | 1.1228 | 0.7326 | 0.3853 |
| LogReg | Raw | 0.9057 | 0.2283 | 0.4761 | 0.3591 | 0.9868 | 0.7291 | 0.3709 |
| LogReg | Isotonic | 0.9051 | 0.2291 | 0.4592 | 0.3602 | 0.9674 | 0.7374 | 0.3759 |
| LogReg | Platt | 0.9057 | 0.2283 | 0.4761 | 0.3591 | 0.9658 | 0.7306 | 0.3724 |
| LogReg | Temperature | 0.9057 | 0.2283 | 0.4761 | 0.3591 | 0.9892 | 0.7291 | 0.3706 |
| SVM | Raw | 0.9021 | 0.2258 | 0.4910 | 0.3550 | 7.1622 | 0.5000 | -9.1719 |
| SVM | Isotonic | 0.9008 | 0.2249 | 0.4857 | 0.3540 | 1.0022 | 0.7465 | 0.3859 |
| SVM | Platt | 0.9021 | 0.2258 | 0.4910 | 0.3550 | 1.0039 | 0.7398 | 0.3936 |
| SVM | Temperature | 0.9021 | 0.2258 | 0.4910 | 0.3550 | 1.4026 | 0.7366 | 0.3889 |
| XGB | Raw | 0.9131 | 0.2503 | 0.4910 | 0.3705 | 1.0167 | 0.7468 | 0.3917 |
| XGB | Isotonic | 0.9116 | 0.2521 | 0.4825 | 0.3715 | 0.9999 | 0.7429 | 0.3897 |
| XGB | Platt | 0.9131 | 0.2503 | 0.4910 | 0.3705 | 1.0175 | 0.7483 | 0.3924 |
| XGB | Temperature | 0.9131 | 0.2503 | 0.4910 | 0.3705 | 1.0255 | 0.7484 | 0.3935 |

**Supplementary Figure S1 |** Beeswarm plots of top features for (a) XGBoost, pre-ablation; (b) ANN, pre-ablation; and (c) ANN, post-ablation.

Each point represents an individual in the held-out test set; color reflects the min-max normalized feature value (blue = lower, red = higher), and horizontal position reflects each feature's contribution to that individual's predicted risk. Features are ordered by mean absolute SHAP value. Asterisked features are binary missingness indicators. Dx = diagnosis; ED = emergency department. Feature attributions were computed via TreeSHAP for XGBoost (a), which computes exact Shapley values (Lundberg et al., 2020), and via GradientShap for ANN (b, c), which approximates attributions by sampling along paths from a set of reference baseline values (Erion et al., 2021); this difference in method is why ANN's attributions show a more granular, clustered distribution than XGBoost's exact TreeSHAP values. See Supplementary Table S3 for full feature definitions.

(a) **XGBoost, pre-ablation**

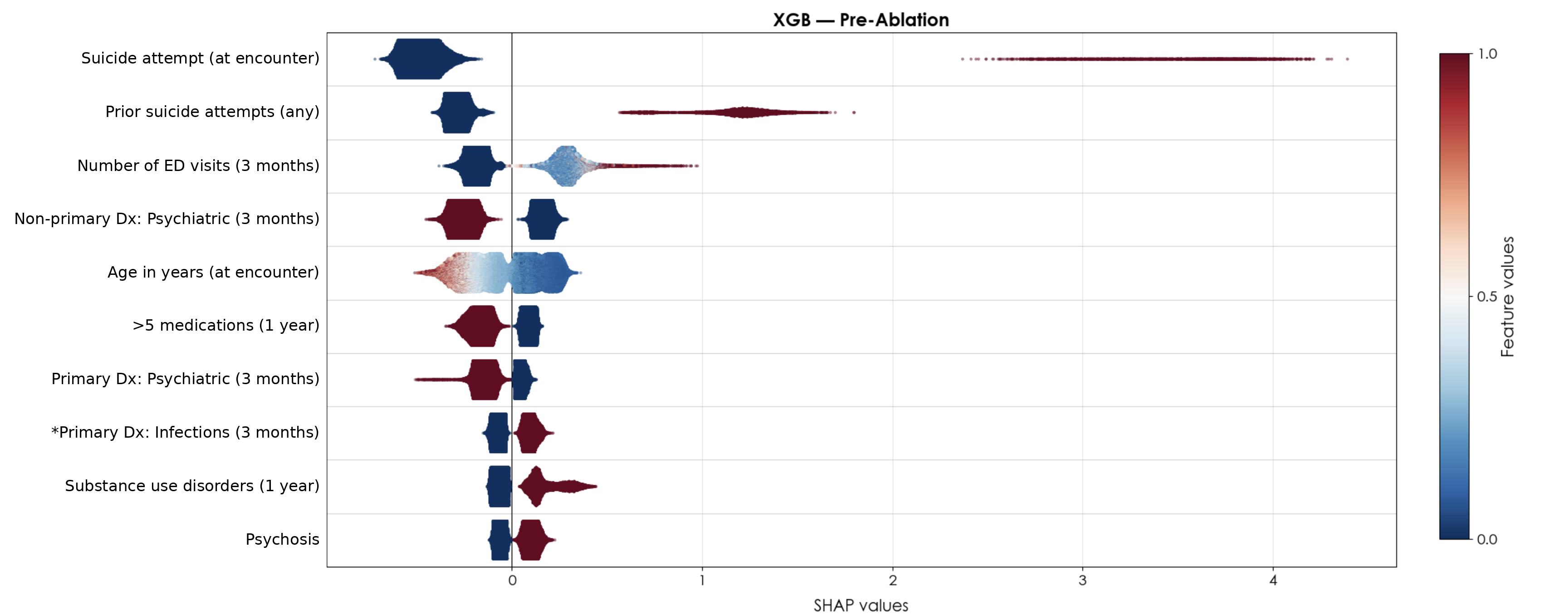

(b) **ANN, pre-ablation**

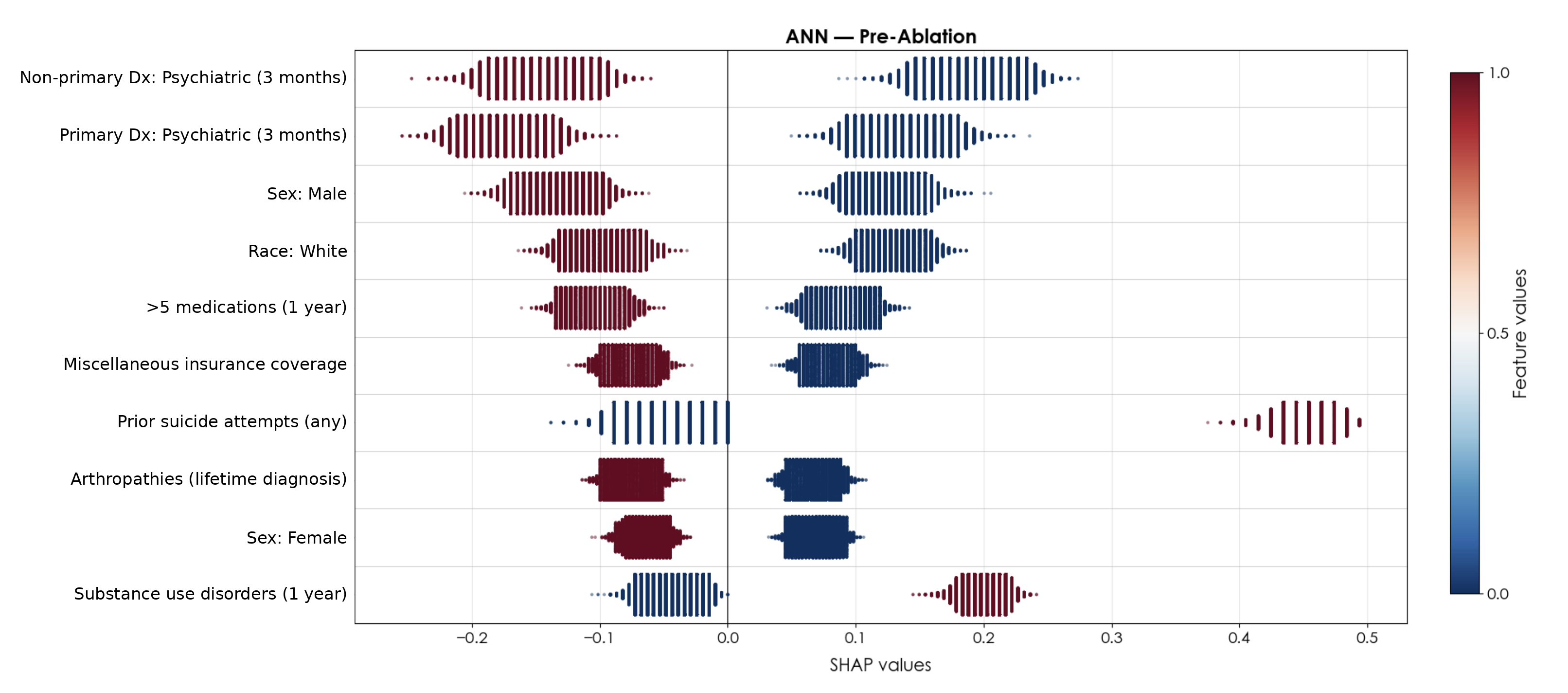

(c) **ANN, post-ablation**

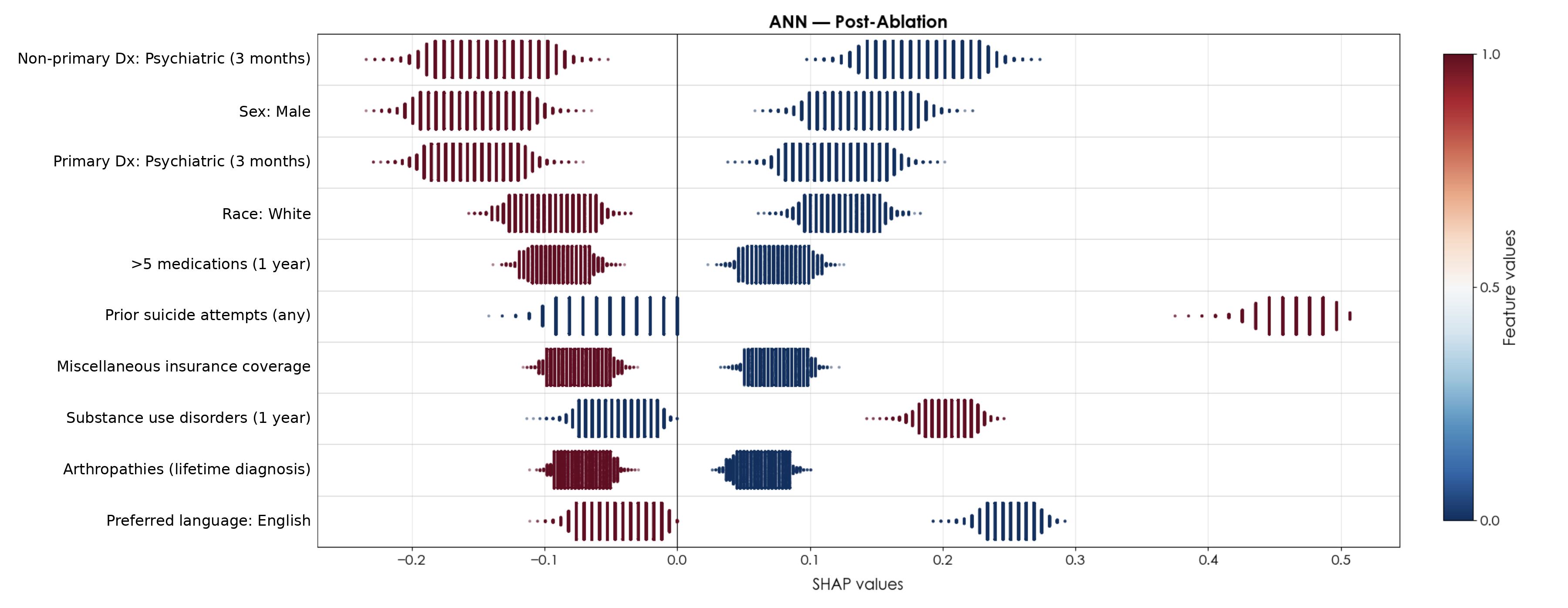
